# Deep Learning of Left Atrial Structure and Function Provides Link to Atrial Fibrillation Risk

**DOI:** 10.1101/2021.08.02.21261481

**Authors:** James P. Pirruccello, Paolo Di Achille, Seung Hoan Choi, Shaan Khurshid, Mahan Nekoui, Sean J. Jurgens, Victor Nauffal, Kenney Ng, Samuel F. Friedman, Kathryn L. Lunetta, Anthony A. Philippakis, Jennifer E. Ho, Steven A. Lubitz, Patrick T. Ellinor

**Author notes:** **Corresponding Author:** James P. Pirruccello, MD, Cardiology Division, Massachusetts General Hospital, 185 Cambridge Street, CPZN / 3.187, Boston, MA 02114.

## Abstract

**Aims:** Increased left atrial (LA) volume is a known risk factor for atrial fibrillation (AF). There is also emerging evidence that alterations in LA function due to an atrial cardiomyopathy are associated with an increased risk of AF. The availability of large-scale cardiac MRI data paired with genetic data provides a unique opportunity to assess the joint genetic contributions of LA structure and function to AF risk.

**Methods and results:** We developed deep learning models to measure LA traits from cardiovascular magnetic resonance imaging (MRI) in 40,558 UK Biobank participants and integrated these data to estimate LA minimum (LAmin), maximum (LAmax), and stroke volume (LASV), as well as emptying fraction (LAEF). We conducted a genome-wide association study (GWAS) in 35,049 participants without pre-existing cardiovascular disease, identifying 20 common genetic loci associated with LA traits. Eight of the loci associated with LA traits were previously associated with AF: the AF risk alleles were associated with an increased LA minimum volume (LAmin) and a decreased LAEF. A Mendelian randomization analysis confirmed that AF causally affects LA volume (IVW P = 6.2E-06), and provided evidence that LAmin causally affects AF risk (IVW P = 4.7E-05). In UK Biobank participants, a polygenic prediction of LAmin was significantly associated with risk for AF (HR 1.09 per SD; P = 1.6E-36) and ischemic stroke (HR 1.04 per SD; P = 4.7E-03).

**Conclusions:** We performed the largest and highest resolution assessment of LA structure and function to date. We then identified 20 common genetic variants associated with LA volumes or LAEF, 19 of which were novel. We found that a polygenic prediction of the minimal LA volume was associated with AF and stroke. Finally, we found an inverse relation between genetic variants associated with AF risk and LAEF. Our findings provide evidence of a causal relation between LA contractile function and AF.

## Introduction

Atrial fibrillation (AF) is a common arrhythmia that is projected to affect up to 12 million Americans by 2050^1^. As a leading cause of stroke^2,3^, the risk factors for AF have been the subject of extensive investigation^4–6^. Enlargement of left atrial (LA) volumes is commonly observed with hypertension^7^, heart failure^8^, or after a diagnosis of AF^9,10^—and AF plays a causal role in this process^11^. Enlargement of the LA and decreased LA function have also been identified as independent risk factors for AF^10,12–17^ and stroke^18–20^. Together these atrial structural, contractile, or electrophysiological changes that have clinical consequences have been termed atrial cardiomyopathies^21,22^.

The link between LA function and AF risk has prompted interest in determining the heritability and common genetic basis for variation in LA measurements. A large-scale genome-wide association study (GWAS) in 30,201 individuals with LA measurements ascertained by echocardiography did not identify any loci with P < 5E-08^23^. Recently, a genome-wide association study of deep learning-derived diastolic measurements in 34,245 UK Biobank participants identified one variant associated with LA volume near *NPR3*^24,25^.

Taking advantage of the precision of cardiovascular magnetic resonance imaging (MRI), we developed deep learning models to produce two-dimensional measurements of the LA in 40,558 participants in the UK Biobank^26,27^, and applied a surface reconstruction technique to integrate these data into three-dimensional LA volume estimates. We reproduced prior observational associations between LA measurements and AF, heart failure, hypertension, and stroke. We then undertook analyses to identify common genetic variants associated with LA volumes in over 35,000 UK Biobank participants. Finally, using common genetic variants as instruments for Mendelian randomization, we performed bidirectional causal analyses between LA volume and AF.

## Results

### Reconstruction of LA volumes from cardiovascular magnetic resonance images

We trained deep learning models to annotate the LA and left ventricular blood pools in four views (distinct models for the short axis view, and the two-, three-, and four-chamber long axis views). We then applied these models to all available UK Biobank cardiovascular magnetic resonance imaging (MRI) data (**Online Methods**)^26–28^. The quality of the deep learning models for measuring the LA was high for the long axis views, and as expected, lower for the short axis views because this view was not designed to capture the LA (**Supplementary Note**). We integrated the data from these separate cross-sections to compute the surface of a 3-dimensional representation of the LA, yielding LA volume estimates at 50 timepoints throughout the cardiac cycle for 40,558 participants (**Figure 1**). We conducted analyses on the maximum LA volume (LAmax), the minimum LA volume (LAmin), the difference between those two volumes (stroke volume; LASV), and the emptying fraction (LASV/LAmax; LAEF).

**Figure 1.**
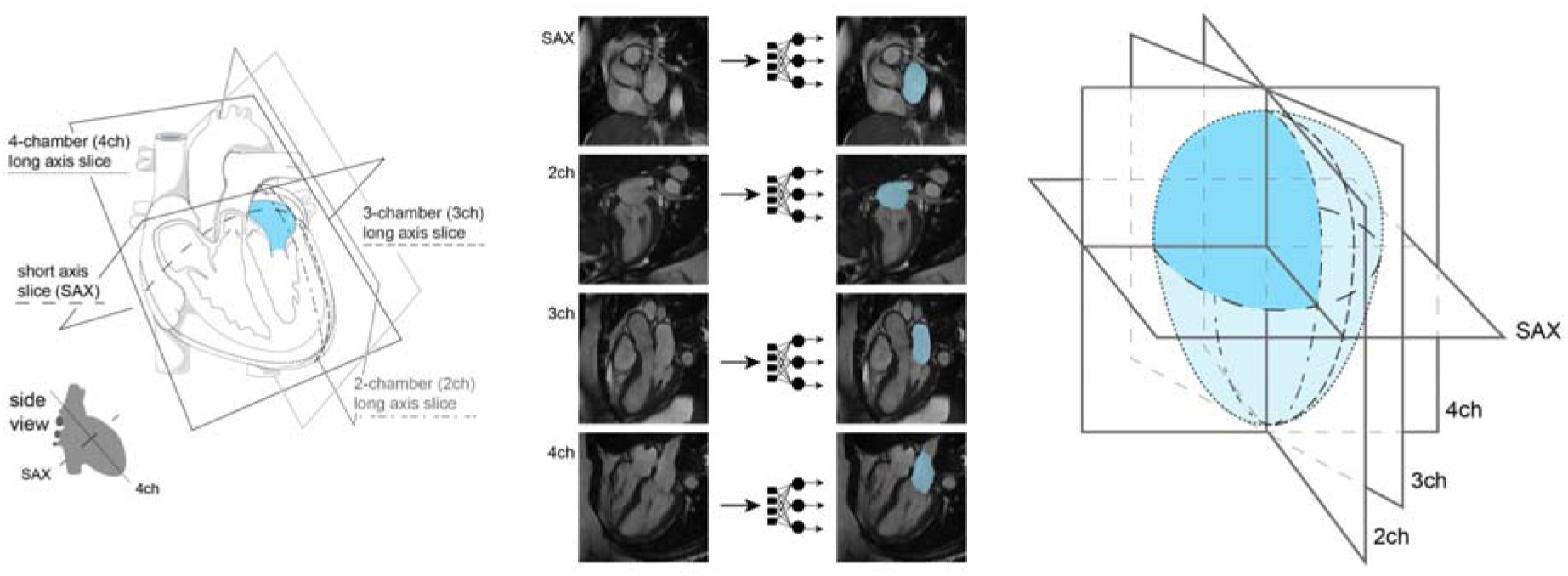
Study overview. **Left panel**: Orientation of the different planes in which images of the atrium were captured. **Middle panel**: Example images from each of the four imaging planes; after interpretation with the deep learning model, the left atrium is colored in blue. **Right panel**: schematic overview representing reconstruction of the left atrium based on information obtained from the deep learning output from the four imaging planes.

### LA traits are associated with AF, heart failure and stroke

We analyzed the pattern of cardiac chamber volumes throughout the cardiac cycle in order to identify individuals with abnormal atrial contraction (**Supplementary Note**). Interestingly, a subset of 1,013 participants with abnormal atrial contraction had markedly elevated LA volumes, similar to those with pre-existing AF (**Figure 2**), and were excluded from downstream analyses.

**Figure 2.**
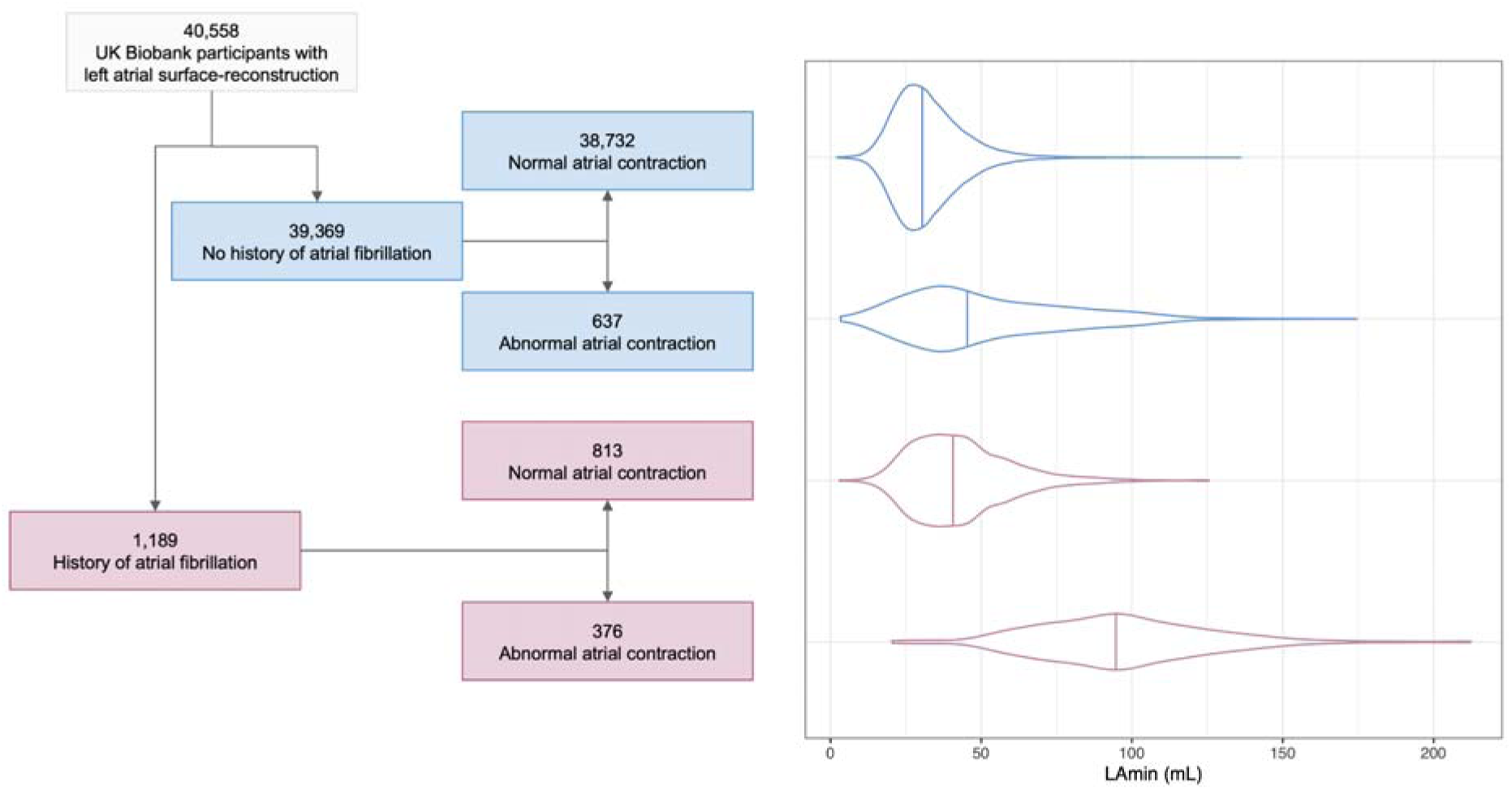
In the **left panel**, a flow diagram breaks down the imaged population into groups with and without AF, and then further into groups that do and do not appear to have normal atrial contraction patterns. In the **right panel**, the LAmin volume is depicted for these groups with violin plots; the median for each group is demarcated with a vertical line.

In the remaining 39,545 participants, we evaluated the association between LA measurements and prevalent or incident AF (**Supplementary Note**). The LA phenotype most strongly associated with AF was the LA minimal volume (LAmin). The 813 individuals with pre-existing AF had a greater LAmin (+8.8mL, P = 9.2E-117). In the ∼2.2 years of follow-up time available on average after MRI acquisition, the risk of incident AF was increased among those with greater LAmin (293 cases; HR 1.73 per standard deviation [SD] increase; 95% CI 1.60-1.88; P = 4.0E-39). We also observed significant associations between LA measurements and hypertension, heart failure, and stroke (**Figure 3** and **Supplementary Tables 1-3**).

**Figure 3.**
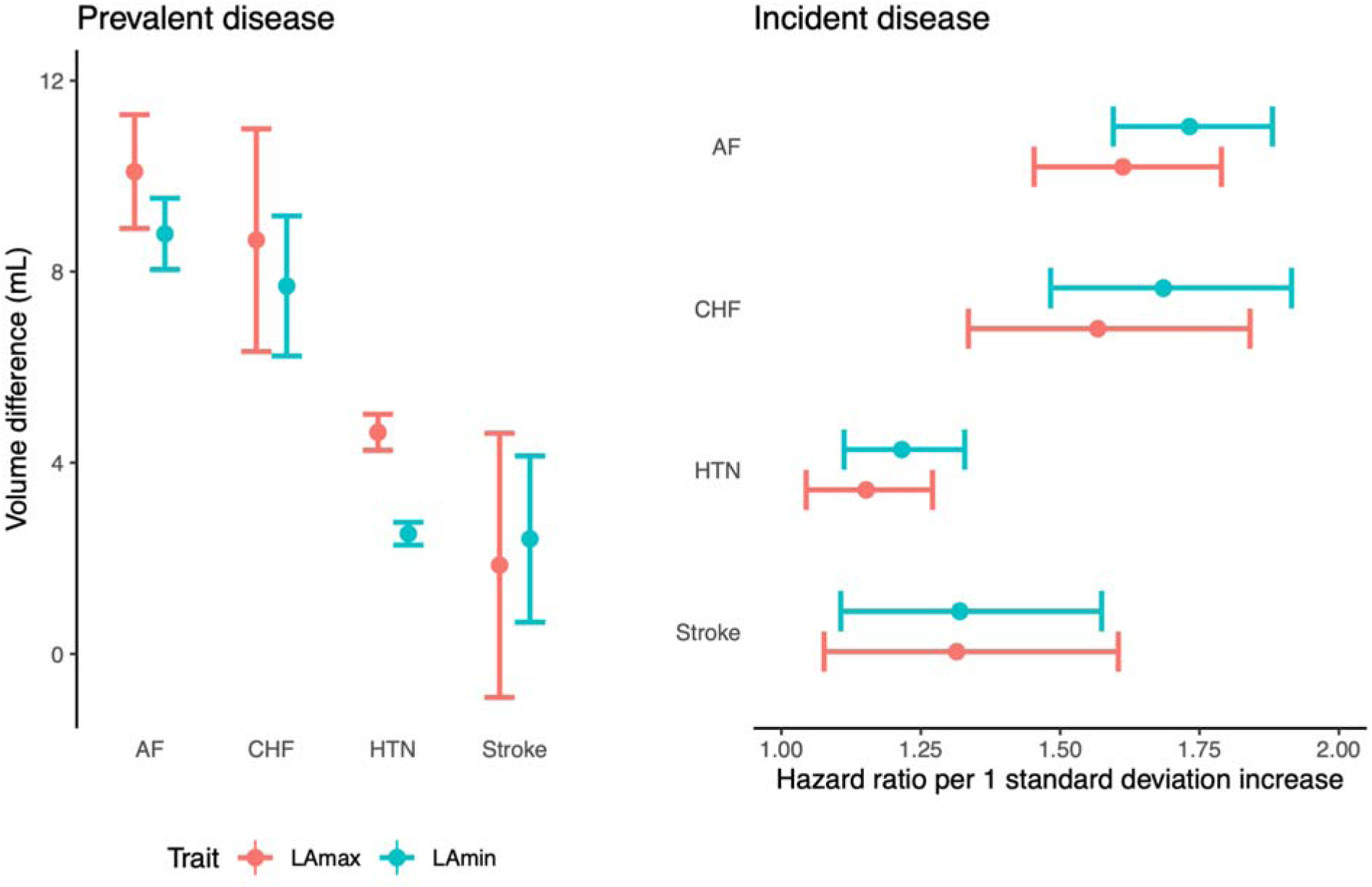
**Left panel** (“Prevalent disease”): the difference in LA volumes (Y axis) between UK Biobank participants with atrial fibrillation (“AF”), heart failure (“CHF”), hypertension (“HTN”), or stroke occurring prior to MRI compared to participants without disease (X axis). **Right panel** (“Incident disease”): hazard ratios for incidence of AF, CHF, HTN, and stroke (Y axis) occurring after MRI per 1 standard deviation increase in LA volumes (X axis). Point estimates are represented by a circle; 95% confidence intervals for the estimate are represented by error bars.

### Common genetic variant analysis of LA size and function identifies 20 loci

After establishing that the LA measurements replicated previously established clinical associations, we then examined the association between common genetic variants and seven LA traits. We conducted these analyses in 35,049 participants with genetic data and without a history of AF, coronary artery disease, or heart failure (**Table 1**; **Supplementary Figure 1**). First, we examined the SNP-heritability of the LA traits which ranged from 0.17 (LAEF) to 0.37 (LAmax; **Supplementary Table 4**). Genetic correlation between the LA measurements ranged from 0.23 (between LAmax and LAEF) to 0.85 (between LAmax and LAmin; **Supplementary Table 4**).

**Table 1:** Participant characteristics.

|  | Women | Men | Both |
| --- | --- | --- | --- |
| **N** | 18,916 | 16,133 | 35,049 |
| **Age at time of MRI** | 64 (8) | 65 (8) | 64 (8) |
| **BMI (kg/m<sup>2</sup>)** | 26 (5) | 27 (4) | 26 (4) |
| **Height (cm)** | 163 (6) | 176 (7) | 169 (9) |
| **Weight (kg)** | 69 (13) | 83 (13) | 75 (15) |
| **Systolic Blood Pressure (mmHg)** | 136 (19) | 142 (17) | 139 (19) |
| **Diastolic Blood Pressure (mmHg)** | 77 (10) | 81 (10) | 79 (10) |
| **Left atrium maximum volume (cm<sup>3</sup>)** | 64 (15) | 79 (19) | 71 (18) |
| **Left atrium minimum volume (cm<sup>3</sup>)** | 28 (9) | 37 (12) | 32 (11) |
| **Left atrium stroke volume (cm<sup>3</sup>)** | 36 (8) | 43 (11) | 39 (10) |
| **Left atrium emptying fraction (%)** | 57 (8) | 54 (7) | 56 (8) |

3 Characteristics of the participants who contributed to the GWAS are listed as mean (standard deviation).

Next, we performed genome-wide association studies (GWAS) for LAmax, LAmin, LAEF, and LASV, as well as for body surface area (BSA)-indexed LA volumes (**Table 2**). Finally, as a sensitivity analysis, we performed GWAS of LA volumes after indexing on left ventricular end diastolic volume (**Supplementary Materials** and **Supplementary Figure 2**). For all analyses, linkage disequilibrium score regression intercepts were near 1, indicating no significant evidence of inflation due to population stratification (**Supplementary Table 5**)^29^.

**Table 2:** GWAS lead SNPs.

| Trait | CHR | BP | dbSNP | Effect Allele | Other Allele | EAF | BETA | SE | P | Nearest Gene |
| --- | --- | --- | --- | --- | --- | --- | --- | --- | --- | --- |
| LAmax | 6 | 31294375 | rs9265346 | G | A | 0.33 | 0.045 | 0.007 | 1.10E-09 | HLA-B |
| LAmax | 6 | 79554193 | rs6926537 | T | A | 0.512 | -0.039 | 0.007 | 9.10E-09 | IRAK1BP1 |
| LAmax | 6 | 107433645 | rs60237682 | T | C | 0.683 | -0.041 | 0.007 | 3.80E-08 | BEND3 |
| LAmax | 12 | 66343400 | rs1038196 | G | C | 0.486 | 0.039 | 0.007 | 1.10E-08 | HMGA2 |
| LAmax | 19 | 46213416 | rs62111731 | A | G | 0.479 | 0.042 | 0.007 | 7.10E-10 | FBXO46 |
| LAmax indexed | 2 | 179650954 | rs6715901 | G | A | 0.507 | -0.043 | 0.008 | 1.20E-08 | TTN |
| LAmax indexed | 4 | 111665783 | rs2634073 | T | C | 0.166 | 0.057 | 0.01 | 2.00E-08 | PITX2 |
| LAmax indexed | 5 | 32831670 | rs13154066 | T | C | 0.402 | -0.05 | 0.008 | 6.60E-11 | NPR3 |
| LAmax indexed | 6 | 31229203 | rs9264391 | G | A | 0.893 | -0.089 | 0.014 | 9.30E-11 | HLA-C |
| LAmax indexed | 6 | 107442277 | rs9480737 | A | G | 0.682 | -0.05 | 0.008 | 8.80E-10 | BEND3 |
| LAmax indexed | 19 | 46166806 | rs140153691 | T | TGC | 0.639 | -0.047 | 0.008 | 4.10E-09 | GIPR |
| LAmax indexed | 22 | 26156505 | rs133873 | T | A | 0.775 | -0.055 | 0.009 | 9.60E-10 | MYO18B |
| LAEF | 1 | 51322205 | rs79948214 | A | G | 0.984 | 0.161 | 0.029 | 4.70E-08 | FAF1 |
| LAEF | 1 | 116310967 | rs4074536 | T | C | 0.702 | -0.052 | 0.008 | 7.80E-11 | CASQ2 |
| LAEF | 14 | 23874117 | rs440466 | T | C | 0.622 | 0.056 | 0.008 | 7.30E-14 | MYH6 |
| LAEF | 22 | 26159289 | rs133885 | G | A | 0.548 | 0.046 | 0.007 | 3.60E-10 | MYO18B |
| LAmin | 6 | 31294375 | rs9265346 | G | A | 0.33 | 0.044 | 0.007 | 8.30E-09 | HLA-B |
| LAmin | 6 | 79518638 | rs35790661 | C | CCA | 0.63 | -0.043 | 0.007 | 4.40E-09 | IRAK1BP1 |
| LAmin | 6 | 107442277 | rs9480737 | A | G | 0.682 | -0.041 | 0.007 | 3.10E-08 | BEND3 |
| LAmin | 10 | 92681480 | rs780162510 | CATA | C | 0.49 | 0.04 | 0.007 | 2.20E-08 | ANKRD1 |
| LAmin | 11 | 65336819 | rs3782089 | C | T | 0.928 | 0.074 | 0.013 | 4.70E-08 | SSSCA1 |
| LAmin | 15 | 99248018 | rs4966014 | C | T | 0.3 | 0.045 | 0.008 | 4.20E-09 | IGF1R |
| LAmin | 19 | 46315357 |  | AT | A | 0.476 | 0.047 | 0.007 | 1.20E-11 | RSPH6A |
| LAmin | 22 | 26164079 | rs133902 | C | T | 0.56 | -0.041 | 0.007 | 4.30E-09 | MYO18B |
| LAmin indexed | 2 | 3912741 | rs56289263 | C | T | 0.938 | -0.093 | 0.017 | 4.40E-08 | DCDC2C |
| LAmin indexed | 2 | 179650954 | rs6715901 | G | A | 0.507 | -0.041 | 0.007 | 4.50E-08 | TTN |
| LAmin indexed | 4 | 24289655 | rs1533093 | C | T | 0.862 | -0.062 | 0.011 | 3.80E-08 | DHX15 |
| LAmin indexed | 4 | 111665783 | rs2634073 | T | C | 0.166 | 0.057 | 0.01 | 1.20E-08 | PITX2 |
| LAmin indexed | 5 | 32831670 | rs13154066 | T | C | 0.402 | -0.043 | 0.008 | 1.40E-08 | NPR3 |
| LAmin indexed | 6 | 31294375 | rs9265346 | G | A | 0.33 | 0.047 | 0.008 | 1.00E-08 | HLA-B |
| LAmin indexed | 6 | 107442277 | rs9480737 | A | G | 0.682 | -0.049 | 0.008 | 1.00E-09 | BEND3 |
| LAmin indexed | 8 | 97223162 | rs35216833 | C | T | 0.456 | 0.042 | 0.007 | 1.00E-08 | UQCRB |
| LAmin indexed | 10 | 92586289 | rs112343361 | A | ACT | 0.497 | -0.046 | 0.008 | 1.80E-09 | HTR7 |
| LAmin indexed | 15 | 99248018 | rs4966014 | C | T | 0.3 | 0.047 | 0.008 | 1.80E-08 | IGF1R |
| LAmin indexed | 17 | 45097337 | rs8078336 | G | T | 0.965 | -0.137 | 0.025 | 2.70E-08 | GOSR2 |
| LAmin indexed | 19 | 46292259 | rs7246377 | G | A | 0.534 | -0.049 | 0.007 | 2.80E-11 | DMWD |
| LAmin indexed | 22 | 26156512 | rs133874 | A | G | 0.773 | -0.061 | 0.009 | 6.80E-12 | MYO18B |
| LASV | 6 | 31225196 | rs199610865 | T | TA | 0.853 | -0.055 | 0.01 | 2.90E-08 | HLA-C |
| LASV | 14 | 23869029 | rs376439 | A | G | 0.606 | 0.045 | 0.007 | 2.70E-10 | MYH6 |
| LASV indexed | 6 | 31225196 | rs199610865 | T | TA | 0.853 | -0.067 | 0.011 | 4.70E-10 | HLA-C |
| LASV indexed | 9 | 136138765 | rs8176685 | GCGCC<br>CACCA<br>CTA | G | 0.82 | -0.059 | 0.01 | 3.20E-09 | OBP2B |
| LASV indexed | 14 | 23869029 | rs376439 | A | G | 0.606 | 0.048 | 0.008 | 8.10E-10 | MYH6 |

1 **BP:** GRCh37-base position. **dbSNP:** dbSNP identifier, where available. **EAF:** Effect allele frequency.
2 **BETA:** BOLT-LMM effect size of the effect allele. **SE:** Standard error. **P:** BOLT-LMM P-value.
3 “Indexed” indicates that the trait has been divided by body surface area.

In the GWAS of LA traits conducted without indexing to BSA, we identified five loci associated with LAmax, eight with LAmin, four with LAEF, and two with LASV (**Figure 4**). Four loci were shared between LAmax and LAmin, with lead SNPs near *HLA-B, IRAK1BP1, BEND3*, and *FBXO32*/*RSPH6A*. LAmax was additionally associated with SNPs at the *HMGA2* locus, and LAmin was associated with SNPs near *ANKRD1, SSSCA1, IGF1R*, and *MYO18B*. The four LAEF loci were located near *FAF1, CASQ2, MYH6*, and *MYO18B*. The two LASV-associated loci included SNPs near *HLA-C* and *MYH6*. Indexing on BSA yielded three additional loci shared by both LAmax and LAmin (*TTN, PITX2*, and *NPR3*), as well as *MYO18B* for LAmax, *UQCRB, HTR7*, and *GOSR2* for LAmin, and *OBP2B* for LASV. Additional loci were identified in a sensitivity analysis that accounted for left ventricular end diastolic volume (LVEDV; **Supplementary Table 6**). Because adjustment for heritable covariates can induce spurious association signals, interpretation of these loci requires caution (see **Supplementary Note**)^30^.

**Figure 4.**
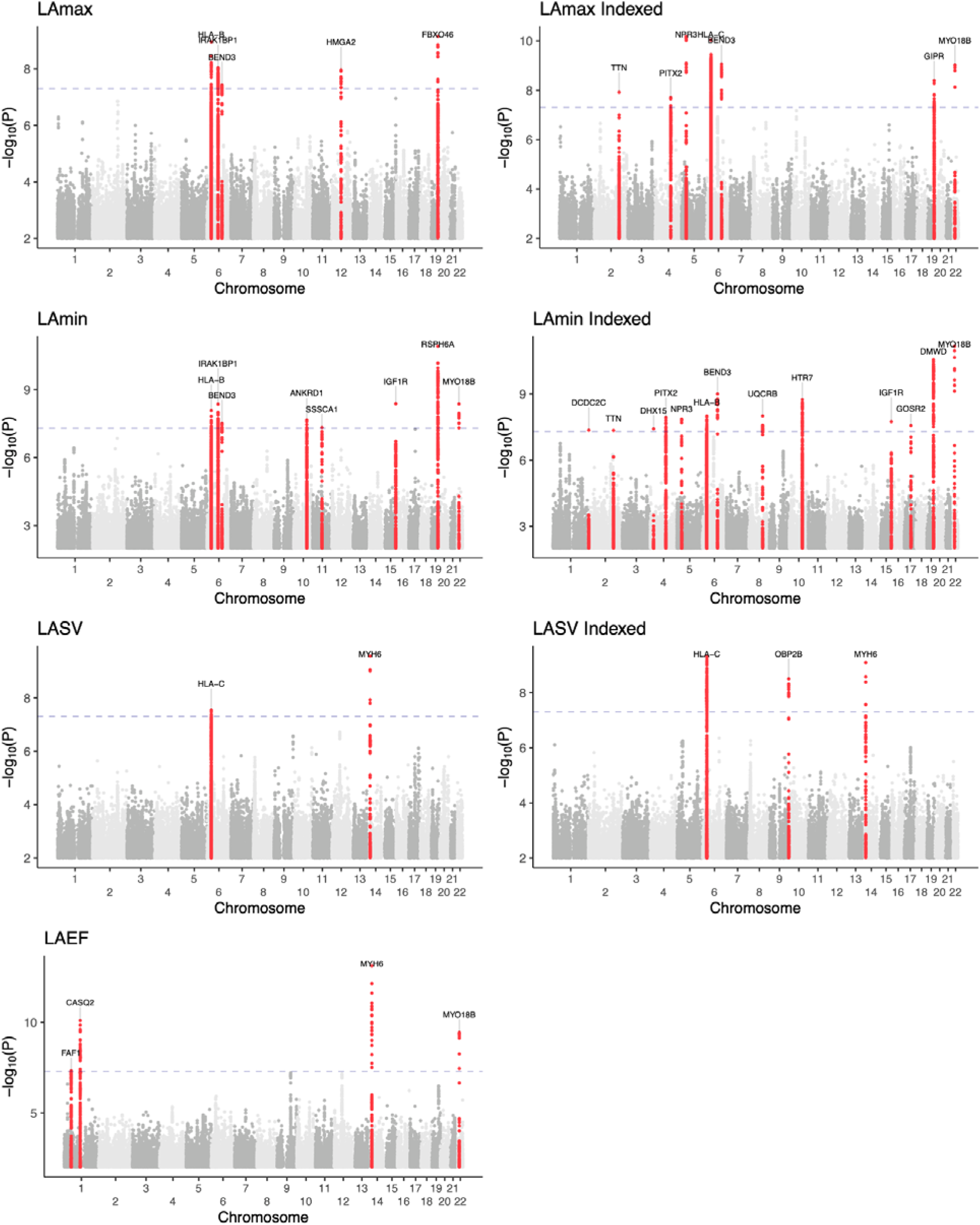
Manhattan plots show the chromosomal position (**X-axis**) and the strength of association (-log10 of the P value, **Y-axis**) for all raw and BSA-indexed phenotypes. Loci that contain SNPs with P < 5E-08 are colored red and labeled with the name of the nearest gene to the most strongly associated SNP.

### Genetic relationship between AF risk and LA dysfunction

To gain more insight into the genetic relationship between LA measurements and AF, we first evaluated their genetic correlations. Using *ldsc*, the strongest genetic correlation was found between LAmin and AF (rg 0.37, P = 2.0E-10), a direction of effect that corresponds to a positive correlation between LA dysfunction (i.e., increased LAmin) and risk for AF (**Supplementary Table 7**)^31,32^. We also tested for association between LA measurements and stroke (all-cause or cardioembolic) from MEGASTROKE; the strongest association was between LAmin and all-cause stroke with nominal significance (rg 0.21, P = 0.01), which was directionally concordant with increased AF risk^33^.

We then assessed the overlap between the 20 distinct LA loci identified in our study and 134 loci previously found to be associated with AF^32^. We found that 8 of the 20 LA loci overlapped with an AF locus, which was a significant enrichment based on permutation testing (P = 2E-04; minimum possible P = 1E-04, see Methods)^34^. The 8 loci found in both the LA GWAS and the AF GWAS are nearest to *FAF1/C1orf85, CASQ2, TTN, PITX2, MYH6/MYH7, IGF1R, GOSR2*, and *MYO18B*. At all 8 loci, the effect of each SNP on AF risk was in opposition to its effect on LAEF, and in most cases the effect of each SNP on AF was concordant with its effect on LAmin (**Figure 5**). None of the loci that were linked with both LA measurements and AF were associated at genome-wide significance with LAmax.

**Figure 5.**
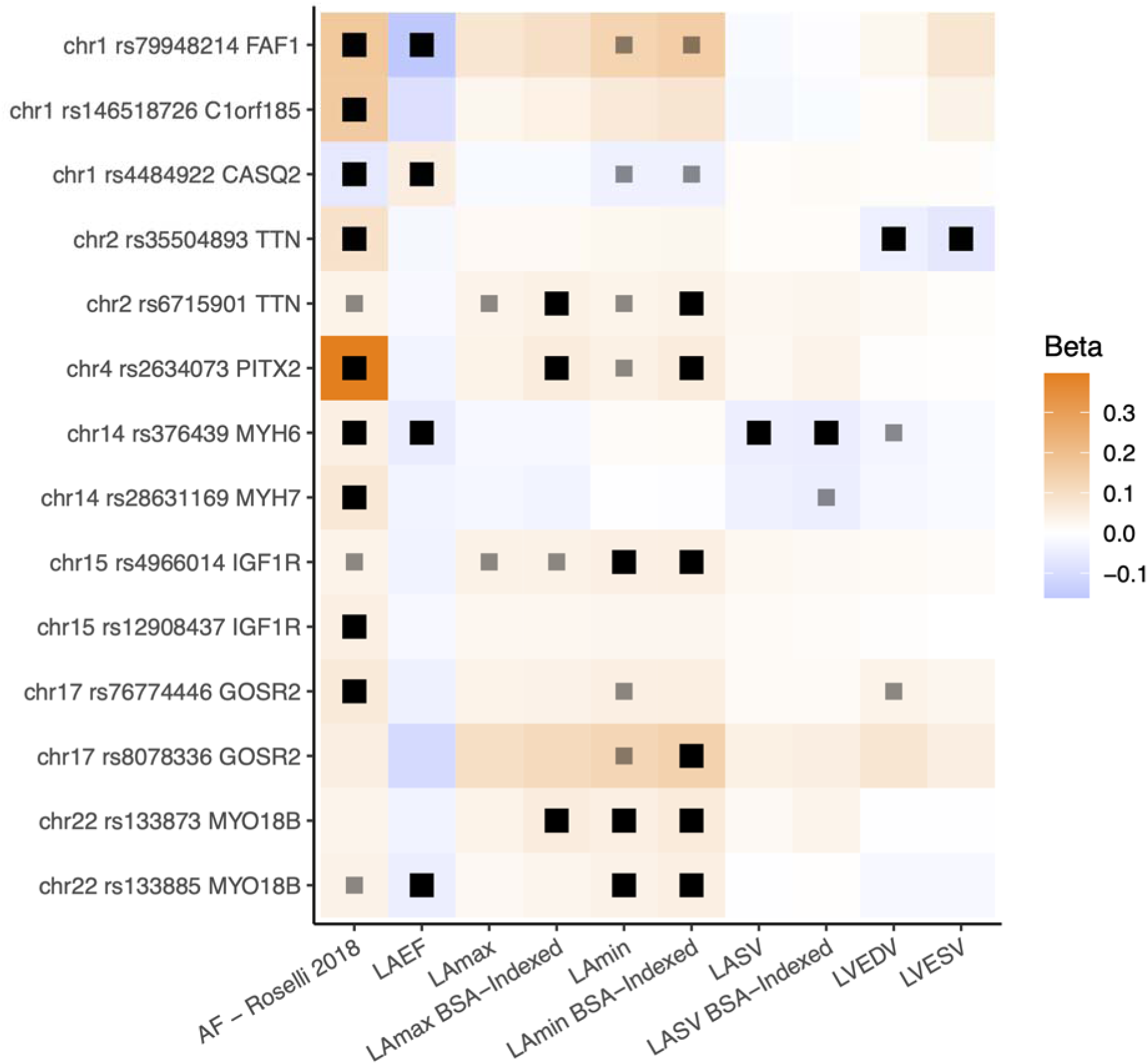
The 8 loci associated with LA measurements and AF are displayed. All loci (except those near *CASQ2* and *PITX2*) have multiple patterns of linkage disequilibrium and are therefore represented multiple times. Black dots represent an association P < 5E-8; gray dots represent P < 5E-6. Effect sizes are oriented with respect to the minor allele. Effect size for AF loci represents the logarithm of the odds ratio.

### Causal link between LA minimal volume and AF risk

Because the genetic correlation analysis suggested that the strongest cross-trait association was between LAmin and AF, we performed bidirectional Mendelian randomization (MR) analyses to assess whether this relationship was causal. First, we assessed the causal effects of LAmin on the risk for AF. Variants that were associated with LAmin with P < 1E-06 were clumped and ambiguous alleles were excluded, leaving 19 SNPs. These variants were looked up in summary statistics from a prior AF GWAS without UK Biobank participants to model the outcome^35^. The inverse variance weighted (IVW) model identified a significant association between LAmin and AF (OR 1.77 per SD increase in LAmin, 95% CI 1.3-2.3, P = 4.7E-05). Simple median, weighted median, MR Egger, and MR-PRESSO showed the same direction of effects (**Supplementary Figure 3**). MR-Egger results did not reach nominal significance, and did not yield evidence for horizontal pleiotropy (intercept P = 0.48). Within the subset of the UK Biobank with imaging data available, three of the 19 SNPs had evidence for pleiotropic association with AF risk factors (**Supplementary Figure 4**) that we derived from the CHARGE-AF risk score^4^. A sensitivity analysis excluding these three variants yielded similar results (IVW OR 1.89 per SD increase in LAmin, P = 7.3E-06; **Supplementary Table 8**; **Supplementary Figure 5**).

We also tested the causal effect of AF on LAmin, with 36 instruments taken from the 2017 AF GWAS, conducted without UK Biobank participants, that were also available in the LAmin summary statistics^35^. Increasing genetic risk of AF was significantly associated with LAmin (0.086 mL increase per unit increase of log of odds of AF liability, 95% CI 0.049-0.123 mL, P = 6.2E-06) using the IVW approach. The simple median, weighted median, MR-Egger, and MR-PRESSO exhibited similar directional effects and a nominal significance. The intercept of the MR-Egger and MR-Egger bootstrap were not significantly different from zero (MR-Egger intercept P = 0.83, MR-Egger bootstrap intercept P = 0.40; **Supplementary Figure 6**).

### A polygenic risk score for AF is associated with LA phenotypes

We constructed a 1.1-million SNP polygenic risk score (PRS) with PRScs from the Christophersen, *et al*, AF GWAS that did not include UK Biobank participants, and applied this score in UK Biobank participants with imaging^35,36^. After excluding participants with a history of AF diagnosed prior to MRI, and participants with abnormal atrial contraction, 36,518 participants remained for analysis. The AF PRS was statistically significantly associated with all measures of LA size and function, with a small effect size (**Supplementary Table 9**). The strongest association was with LAmin (0.049 SD increase in LAmin per SD increase in the PRS; 95% CI 0.039-0.058; P = 9.8E-24).

### A polygenic estimate of LA volume predicts AF, stroke, and heart failure

We created a 1.1 million SNP genome-wide polygenic score for each LA trait using PRScs^36^. We tested this score in the 421,339 UK Biobank participants who did not participate in the LA GWAS, of whom 22,356 developed AF. The strongest association was with the BSA-indexed LAmin polygenic score, which was linked to a modestly increased risk for incident AF or atrial flutter (HR = 1.09 per 1 SD increase in the score; P = 1.6E-36) (**Figure 6**; **Supplementary Table 10**). This score was also associated with small increases in risks of incident all-cause stroke (7,823 cases; HR = 1.04 per SD; P = 4.7E-04), ischemic stroke (5,492 cases; HR = 1.04 per SD; P = 4.7E-03), and heart failure (11,465 cases; HR = 1.05 per SD; P = 2.6E-08). In a sensitivity analysis that censored participants who developed AF prior to a diagnosis of heart failure, the magnitude of effect and strength of association between the LAmin score and heart failure was attenuated (8,247 cases; HR = 1.03 per SD; P = 0.01; **Supplementary Table 11**).

**Figure 6.**
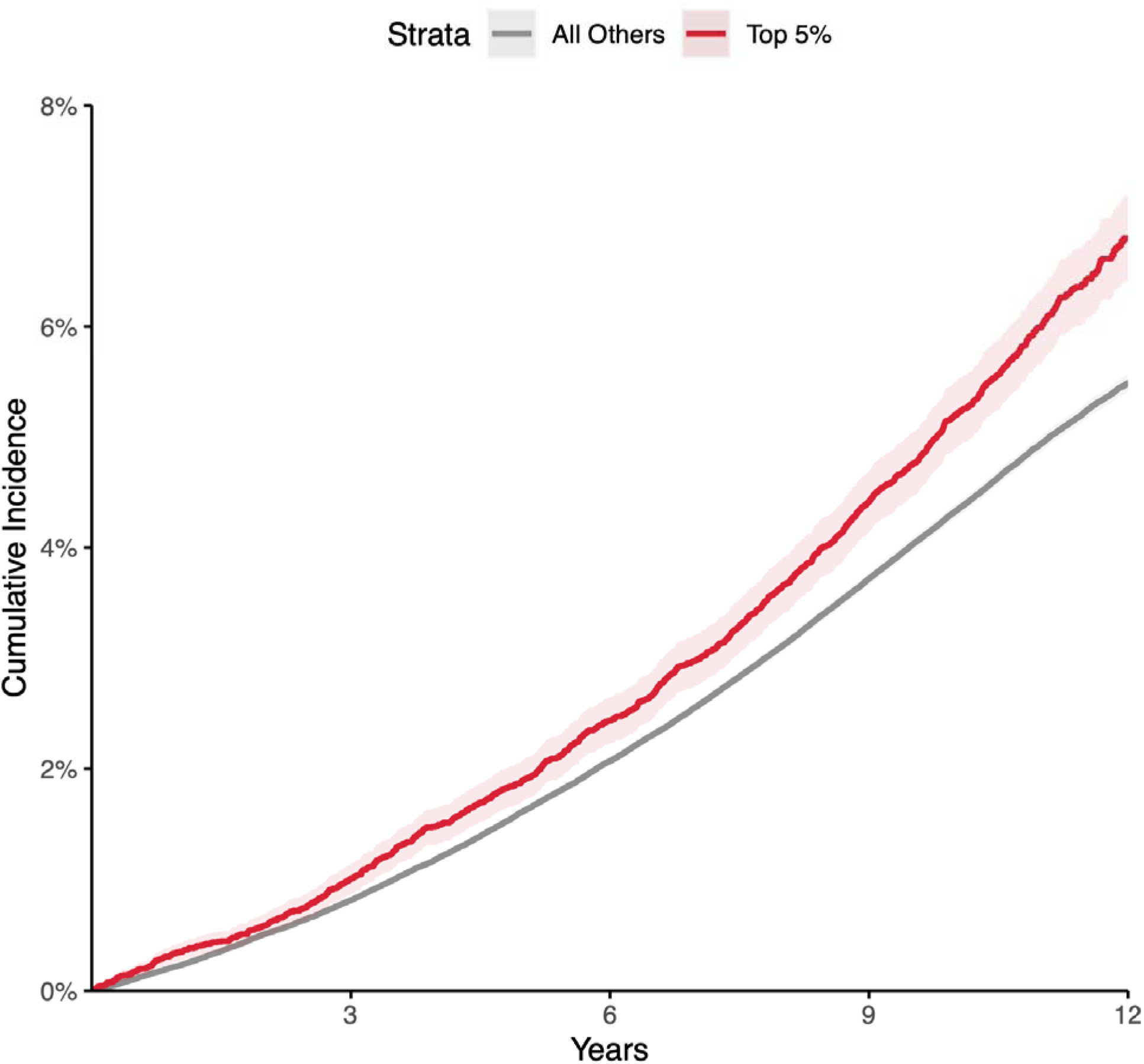
Disease incidence curves for the 417,881 participants who were unrelated to within 3 degrees of the participants who underwent MRI in the UK Biobank. Those in the top 5% for the BSA-indexed LAmin PRS are depicted in red; the remaining 95% are in gray. **X-axis**: years since enrollment in the UK Biobank. **Y-axis**: cumulative incidence of AF (19,875 cases in the bottom 95% and 1,272 cases in the top 5%). Those in the top 5% of genetically predicted LAmin-indexed had an increased risk of AF (Cox HR 1.22, P = 1.2E-11) compared with those in the remaining 95% in up to 12 years of follow-up time after UK Biobank enrollment.

## Discussion

We used a unique resource of more than 40,000 cardiac MRI images available in the UK Biobank to enable one of the largest and highest resolution assessments of LA structure and function to date. We trained deep learning models to segment LA cross sections from cardiovascular MRI data and then derived estimates of LA volume. In turn, we performed an extensive series of epidemiological, genetic, polygenic and Mendelian randomization analyses to link these LA traits to cardiovascular outcomes. Our findings permit at least five primary conclusions.

First, we were able to replicate previous observations demonstrating associations between greater LA volume and cardiovascular diseases^7–10,19,20^. Participants with a history of AF had larger LA volumes; and participants with larger LA volumes were more likely to be subsequently diagnosed with AF, stroke, or heart failure.

Second, these measurements enabled the largest genetic analysis to date of LA measurements. To our knowledge, one locus (near *NPR3*) has previously been associated at genome-wide significance with LA measurements^25^. In this work, 20 distinct genetic loci were associated with LAmax, LAmin, LAEF, LASV, or the BSA-indexed versions of these phenotypes. Forty percent of these loci (8 of 20) were previously associated with AF, significantly more than expected by chance.^32^. At all 8 loci, the allele associated with increased AF risk was directionally associated with a lower LAEF, and generally with greater LA volumes. The uniformly opposed effect directions of these SNPs for AF risk and LAEF may be consistent with the concept of atrial cardiomyopathy^22^.

As an example of the pattern of opposed SNP effects on LAEF and AF risk, we identified a missense variant within *CASQ2* (rs4074536; p.Thr66Ala) as a lead SNP for LAEF on chromosome 1. The T allele of this SNP (encoding Thr66) corresponds with a reduced LAEF in our GWAS, and with reduced expression of *CASQ2* in the right atrial appendage and left ventricle in GTEx^37^. This variant is also in LD (r^2^=1.0) in non-African 1KG populations for the AF lead SNP rs4484922^32,38^. In the study by Roselli and colleagues, the rs4484922-G allele is associated with an increased risk for AF; notably, that risk-increasing allele corresponds to the LAEF-reducing T allele of rs4074536. The rs4074536-T allele has also previously been associated with a longer QRS complex duration^39,40^. *CASQ2* encodes calsequestrin 2, which resides in the sarcoplasmic reticulum in abundance and binds to calcium ions during the cardiac cycle. Missense variants in this gene have also been associated with catecholamine-induced polymorphic ventricular tachycardia, typically following a recessive inheritance pattern^41,42^.

Even among LA-associated loci that were not previously associated with AF, several showed the same consistent pattern of inverse effect between AF risk and LAEF (e.g., near *NPR3, SSSCA1*, and *HMGA2*). However, this pattern did not uniformly hold. For example, at the gene-dense locus near *FBXO46*/*DMWD*/*RPSH6A*, the LA volume-increasing (and LAEF-decreasing) variants were weakly associated with decreased AF risk.

Also notable was the *PITX2* locus, which was the first locus associated with AF. In the present GWAS, SNPs at that locus were associated with BSA-indexed LAmax and LAmin. The lead SNP for AF (rs2129977 from Roselli, *et al*, 2018) was in close LD with the lead SNP for LAmax and LAmin (rs2634073; r^2^ = 0.85)^32,38^. Consistent with clinical expectations, the AF risk allele was associated with greater left atrial maximum and minimum volumes. These analyses excluded participants with a history of AF or abnormal atrial contraction on MRI; therefore, these results support the hypothesis that the *PITX2* locus may be associated with an increase in LA volume that occurs prior to AF onset.

Fourth, we developed polygenic scores to gain additional insight into the relationship between LA volumes and cardiovascular diseases. A genome-wide 1.1-million variant AF PRS derived from Christophersen, *et al*, 2017 was associated with all of the LA phenotypes—and most strongly with LAmin—even after excluding participants known to have AF^35^. This genetic evidence is consistent with and extends prior observational evidence, and suggests that some of the genetic drivers of AF risk may manifest in ways that are detectable in LA size and function.

A 1.1-million variant polygenic predictor of BSA-indexed LAmin was modestly associated with incident AF (**Figure 6**), and weakly with stroke, in the UK Biobank. The score was also associated with heart failure—an association which was almost completely attenuated after excluding participants who were diagnosed with AF prior to heart failure. This attenuation suggests that much of the heart failure association may be mediated through AF.

Finally, we found strong evidence of genetic correlation between LA phenotypes and AF. We pursued Mendelian randomization analyses to more formally assess the hypothesis of bidirectional causation between LA phenotypes and AF. These revealed strong evidence of a causal effect of AF on LAmin, as has been previously observed^11^. There was also evidence that LA volumes, particularly LAmin, may be causal for AF. The causal effect persisted even after excluding three variants associated with at least one risk factor from CHARGE-A^4^. However, because AF can be paroxysmal and remain undiagnosed, we cannot exclude the possibility of cryptic reverse causation: namely, that some participants may have had larger atria because of undiagnosed paroxysmal AF, such that AF itself induced the genetic association with LA volumes.

## Limitations

This study has several limitations. All LA measurements were derived from deep learning models of cardiovascular MRI. Because a complete trans-axial stack of atrial images was not part of the UK Biobank imaging protocol, the LA measurements are estimates that are interpolated from cross sections of the LA. Because contrast protocols were not used during image acquisition, we were not able to ascertain atrial fibrosis. The deep learning models have not been tested outside of the specific devices and imaging protocols used by the UK Biobank and are unlikely to generalize to other data sets without fine tuning. Disease labels were determined by diagnostic and procedural codes; because AF can be paroxysmal and may go undetected, it is likely that a subset of the participants had undiagnosed AF prior to MRI, which would bias causal estimates of the impact of LA volume on disease risk away from the null. The study population was largely composed of people of European ancestries, limiting generalizability of the findings to global populations. The participants who underwent MRI in the UK Biobank tended to be healthier than the remainder of the UK Biobank population, which itself is likely to be healthier than the general population. At present, there is little follow-up time subsequent to the first MRI visit for most UK Biobank participants.

## Conclusions

Measures of LA structure and function are heritable traits that are associated with AF, stroke, and heart failure. Genetic predictors of LA volume are linked to an elevated risk of AF and, to a lesser extent, stroke and heart failure. In future work, it will be interesting to determine if targeting the genes and pathways associated with abnormalities in LA function will be helpful to reduce the risk of AF, heart failure, and stroke.

## Methods

### Study design

Except where otherwise stated, all analyses were conducted in the UK Biobank, which is a richly phenotyped, prospective, population-based cohort that recruited 500,000 participants aged 40-69 years in the UK via mailer from 2006-2010^43^. We analyzed 487,283 participants with genetic data who had not withdrawn consent as of February 2020. Access was provided under application #7089 and approved by the Partners HealthCare institutional review board (protocol 2019P003144).

Statistical analyses were conducted with R version 3.6 (R Foundation for Statistical Computing, Vienna, Austria).

### Cardiovascular magnetic resonance imaging protocols

At the time of this study, the UK Biobank had released images in over 45,000 participants of an imaging substudy that is ongoing^26,27^. Cardiovascular magnetic resonance imaging was performed with 1.5 Tesla scanners (Syngo MR D13 with MAGNETOM Aera scanners; Siemens Healthcare, Erlangen, Germany), and electrocardiographic gating for synchronization^27^. Several cardiac views were obtained. For this study, four views (the long axis two-, three-, and four-chamber views, as well as the short axis view) were used. In these views, balanced steady-state free precession cines, consisting of a series of 50 images throughout the cardiac cycle for each view, were acquired for each participant^27^. For the three long axis views, only one imaging plane was available for each participant, with an imaging plane thickness of 6mm and an average pixel width and height of 1.83mm. For the short axis view, several imaging planes were acquired. Starting at the base of the heart, 8mm-thick imaging planes were acquired with approximately 2mm gaps between each plane, forming a stack perpendicular to the longitudinal axis of the left ventricle to capture the ventricular volume. For the short axis images, the average pixel width and height was 1.86mm.

### Semantic segmentation and quality control

We labeled pixels using a process similar to that described in our prior work evaluating the thoracic aorta^44^. Cardiac structures were manually annotated in images from the short axis view and the two-, three-, and four-chamber long axis views from the UK Biobank by a cardiologist (JPP). To produce the models used in this manuscript, 714 short axis images were chosen, manually segmented, and used to train a deep learning model with PyTorch and fastai v1.0.61^28,45^. The same was done separately with 98 two-chamber images, 66 three-chamber images, and 445 four-chamber images. The models were based on a U-Net-derived architecture constructed with a ResNet34 encoder that was pre-trained on ImageNet^46–49^. The Adam optimizer was used^50^. The models were trained with a cyclic learning rate training policy^51^. 80% of the samples were used to train the model, and 20% were used for validation. Held-out test sets with images that were not used for training or validation were used to assess the final quality of all models.

Four separate models were trained: one for each of the three long axis views, and one for the short axis view. During training, random perturbations of the input images (augmentations) were applied, including affine rotation, zooming, and modification of the brightness and contrast.

For the short axis images, all images were resized initially to 104×104 pixels during the first half of training, and then to 224×224 pixels during the second half of training. The model was trained with a mini-batch size of 16 (with small images) or 8 (with large images). Maximum weight decay was 1E-03. The maximum learning rate was 1E-03, chosen based on the learning rate finder^28,52^. A focal loss function was used (with alpha 0.7 and gamma 0.7), which can improve performance in the case of imbalanced labels^53^. When training with small images, 60% of iterations were permitted to have an increasing learning rate during each epoch, and training was performed over 30 epochs while keeping the weights for all but the final layer frozen. Then, all layers were unfrozen, the learning rate was decreased to 1E-07, and the model was trained for an additional 10 epochs. When training with large images, 30% of iterations were permitted to have an increasing learning rate, and training was done for 30 epochs while keeping all but the final layer frozen. Finally, all layers were unfrozen, the learning rate was decreased to 1E-07, and the model was trained for an additional 10 epochs. The semantic segmentation model training hyperparameters for the two-, three-, and four-chamber long axis images were similar, and are detailed in the **Supplementary Note**.

Each model was applied to all available images from its respective view that were available in the UK Biobank as of November 2020.

### Poisson surface reconstruction

To integrate the output from each of the four models into one LA volume estimate, Poisson surface reconstruction was performed. Among the views included in the UK biobank cardiac MRI dataset, none fully captures the 3-D anatomical structure of the LA. The short axis stack only occasionally included the lower portion of the chamber, while the three long-axis (i.e., two-, three-, and four-chamber) views provided only single-slice cross-sections of the LA at different orientations. To integrate information from the four incomplete MRI views into a consistent 3D representation of the LA anatomy, we followed a procedure similar to Pirruccello et al. (2021)^54^. Briefly, we first co-rotated the MRI views into the same reference system using standard DICOM metadata (i.e., from the Image Position (Patient) [0020,0032] and Image Orientation (Patient) [0020,0037] tags). Then, we applied the Poisson surface reconstruction algorithm^55^ to interpolate 3-D surfaces through the points marking the boundaries of the LA chamber segmentations. In addition to the interpolation point coordinates, the Poisson algorithm requires as input the local normal directions, which constrain the curvature of the reconstructed surface. In our approach, we assumed that the normals lie onto the MRI view planes and are radially oriented outwards from the center of gravity of the LA segmentation. 3D surfaces of the LA were reconstructed for each of the 50 MRI frames captured during the cardiac cycle. At each timepoint, the volume of the LA was computed using routines for triangulated meshes included in the VTK library (Kitware Inc.). From the reconstructed volume traces, we estimated the maximum and minimum LA volumes, as well as LA stroke volume and emptying fraction.

### Identification of abnormal atrial contraction patterns

We sought to identify participants with abnormal atrial contraction patterns at the time of acquisition of the magnetic resonance images. Although the imaging protocol was ECG-gated, the instantaneous ECG signal was not available. Therefore, we used the filling patterns of the atrium and ventricle as markers of normal filling.

To create a training set, we first pulled CINE videos from the 2-, 3-, and 4-chamber long axis views of all participants with a history of AF. A cardiologist (JPP) evaluated whether the videos appeared to represent a typical cardiac cycle including an atrial contraction. A deep learning model was then trained to classify filling patterns as representing a normal atrial contraction or not. Each input channel represented the pixel counts of a cardiac chamber from a different long-axis view, divided by the maximum number of pixels seen for each channel for that participant, over the entire cardiac cycle. This approach prevented the model from accessing information about the absolute size of the chambers, forcing it instead to identify patterns based on relative size differences throughout the cardiac cycle. In total, 8 channels were used as input: four from the 4-chamber long axis images (left atrium, right atrium, left ventricle, right ventricle), two from the 3-chamber long axis images (left atrium, left ventricle), and two from the 2-chamber long axis images (left atrium, left ventricle). Cases were excluded if all 8 channels were not available. Therefore, the shape of the input was 50×8. Training was performed with FastAI version 2.2.5^28^, using the TimeseriesAI library version 0.2.15 (github.com/timeseriesAI/tsai) to train an InceptionTime model^56^. The Ranger optimization function was used with cross entropy loss, and the number of filters in the InceptionTime model was 32, all of which are the software defaults in the TimeseriesAI library. Ranger incorporates RAdam and Lookahead to improve training stability early and later during training, respectively^57,58^. 20% of samples were randomly chosen as the validation set. The model was trained with a batch size of 32. Variable learning rates from 5E-06 to 5E-03 were permitted during training. Training was conducted using the One-Cycle policy for 20 epochs^51,52^.

### Evaluation of the relationship between the left atrium and cardiovascular diseases

We focused on three disease definitions related to LA structure and function: AF or flutter, ischemic stroke, and heart failure (defined below in **Online Methods**). For prevalent disease that was diagnosed prior to the time of imaging, linear models were used to test for an association between each disease (as a binary independent variable) and LA phenotypes (as the dependent variables), adjusting for the MRI serial number, sex, age, and the interaction between sex and age.

For incident disease, participants with pre-existing diagnoses prior to the MRI were excluded from the analysis. A Cox proportional hazards model was used, with survival defined as the time between MRI and either the time of censoring, or disease diagnosis. The model was adjusted for the MRI serial number, sex, age, the interaction between sex and age, the cubic natural spline of height, the cubic natural spline of weight, and the cubic natural spline of BMI. As a sensitivity analysis, adjustment was additionally made for heart rate, P duration, QRS duration, P-Q interval, QTc interval, left ventricular end systolic volume, left ventricular end diastolic volume, and left ventricular ejection fraction.

### Genotyping, imputation, and genetic quality control

UK Biobank samples were genotyped on either the UK BiLEVE or UK Biobank Axiom arrays and imputed into the Haplotype Reference Consortium panel and the UK10K+1000 Genomes panel^59^. Variant positions were keyed to the GRCh37 human genome reference. Genotyped variants with genotyping call rate < 0.95 and imputed variants with INFO score < 0.3 or minor allele frequency <= 0.005 in the analyzed samples were excluded. After variant-level quality control, 11,253,549 imputed variants remained for analysis.

Participants without imputed genetic data, or with a genotyping call rate < 0.98, mismatch between self-reported sex and sex chromosome count, sex chromosome aneuploidy, excessive third-degree relatives, or outliers for heterozygosity were excluded from genetic analysis^59^. Participants were also excluded from genetic analysis if they had a history of AF or flutter, hypertrophic cardiomyopathy, dilated cardiomyopathy, heart failure, myocardial infarction, or coronary artery disease documented prior to the time they underwent cardiovascular magnetic resonance imaging at a UK Biobank assessment center. Our definitions of these diseases in the UK Biobank are provided in **Supplementary Table 12**.

### Genome-wide association study of the left atrium

We analyzed four primary LA phenotypes, as well as LAmax, LAmin, and LASV estimates that were adjusted for BSA or LVEDV. In total, we conducted 10 genome-wide association studies with these traits. Before conducting genetic analyses, a rank-based inverse normal transformation was applied^60^. All traits were adjusted for sex, age at enrollment, age and age^2^ at the time of MRI, the first 10 principal components of ancestry, the genotyping array, and the MRI scanner’s unique identifier.

BOLT-REML v2.3.4 was used to assess the SNP-heritability of the phenotypes, as well as their genetic correlation with one another using the directly genotyped variants in the UK Biobank^61^. Genome-wide association studies for each phenotype were conducted using BOLT-LMM version 2.3.4 to account for cryptic population structure and sample relatedness^61,62^. We used the full autosomal panel of 714,577 directly genotyped SNPs that passed quality control to construct the genetic relationship matrix (GRM), with covariate adjustment as noted above. Associations on the X chromosome were also analyzed, using all autosomal SNPs and X chromosomal SNPs to construct the GRM (N=732,214 SNPs), with the same covariate adjustments and significance threshold as in the autosomal analysis. In this analysis mode, BOLT treats individuals with one X chromosome as having an allelic dosage of 0/2 and those with two X chromosomes as having an allelic dosage of 0/1/2. Variants with association P < 5·10^−8^, a commonly used threshold, were considered to be genome-wide significant.

We identified lead SNPs for each trait. Linkage disequilibrium (LD) clumping was performed with PLINK-1.9^63^ using the same participants used for the GWAS. We outlined a 5-megabase window (--clump-kb 5000) and used a stringent LD threshold (--r2 0.001) in order to account for long LD blocks. With the independently significant clumped SNPs, distinct genomic loci were then defined by starting with the SNP with the strongest P value, excluding other SNPs within 500kb, and iterating until no SNPs remained. Independently significant SNPs that defined each genomic locus are termed the lead SNPs.

No lead SNPs deviated from Hardy-Weinberg equilibrium (HWE) at a threshold of P < 1E-06^63^.

Linkage disequilibrium (LD) score regression analysis was performed using *ldsc* version 1.0.0^29^. With *ldsc*, the genomic control factor (lambda GC) was partitioned into components reflecting polygenicity and inflation, using the software’s defaults.

### Genetic correlation with atrial fibrillation

We used *ldsc* version 1.0.1 to perform cross-trait LD score regression to estimate genetic correlation between the LA measurements, atrial fibrillation (from Roselli, et al, 2018), and all-cause or cardioembolic stroke (from Malik, *et al*, 2018)^31–33^. Summary stats were pre-processed with the *munge_sumstats*.*py* script from *ldsc* 1.0.1 using the default settings, filtering out variants with imputation INFO scores less than 0.9 or minor allele frequencies below 0.01, as well as strand-ambiguous variants.

### Overlap of left atrial loci with atrial fibrillation loci

We identified the gene nearest to SNPs associated with AF from Supplementary Table 16 of Roselli, *et al*^32^. For this exercise, we used each of the 134 SNPs that achieved association P < 5E-8 in the primary GWAS (column ‘I’) or in the meta-analysis (column ‘AD’). We counted the number of AF nearest genes that fell within 500kb of the LA lead SNPs from our study. We used SNPsnap to generate 10,000 sets of SNPs that matched the LA lead SNPs based on parameters including minor allele frequency, SNPs in linkage disequilibrium, distance from the nearest gene, and gene density^34^. We then repeated the same counting procedure for each of the 10,000 synthetic SNPsnap lead SNP lists, to set a neutral expectation for the number of overlapping AF nearest genes based on chance. This allowed us to compute a one-tailed permutation P value (with the most extreme possible P value based on 10,000 randomly chosen sets of SNPs being 1E-04).

### Mendelian randomization

We sought to assess a potential causal relationship between LAmin and AF using Mendelian randomization (MR). We considered LAmin as the exposure and AF as the outcome. The genetic instruments for LAmin were generated using the genome-wide association results from this analysis. The variants from the exposure summary statistics were clumped with P < 1E-06, r^2^ < 0.001, and a radius of 5 megabases using the *TwoSampleMR* package in R^64^. The variants with ambiguous alleles were removed. 19 variants were harmonized with a large AF GWAS that did not include UK Biobank participants^35^. The inverse variance weighted (IVW) method was performed as the primary MR analysis. We also performed simple median, weighted median, MR-Egger, and MR-PRESSO to account for violations of the instrumental variable assumptions. Since MR-Egger provides robust estimates under the InSIDE (Instrument Strength Independent of Direct Effect) assumption, we additionally conducted the MR-Egger bootstrap method to confirm the results from MR-Egger.

To assess risk of pleiotropy of the LA genetic instruments, each SNP was tested for association with risk factors from CHARGE-AF^4^ within the same participants in which the GWAS was conducted. Association between each of the 19 variants and seven risk factors (height, weight, systolic blood pressure, diastolic blood pressure, use of antihypertensive medications [ascertainment described below in **Online Methods**], diagnosis of diabetes, and current smoking) was tested in a linear regression model that accounted for age and age^2^ at the time of MRI, sex, the MRI serial number, the genotyping array, and genetic principal components 1-10. Associations were considered significant if they exceeded Bonferroni significance (P < 3.8E-04).

To understand the bidirectional causal effects, we also performed an MR analysis using AF variants from the 2017 GWAS as the exposure and LAmin as the outcome. After applying the same clumping threshold and filtering methods to AF summary statistics, 36 remaining variants were harmonized with the LAmin association results and used to construct the instrumental variable. The primary and sensitivity analyses were then conducted in the same manner as described above.

### Polygenic risk analysis

A polygenic score for the LAmin GWAS was computed using PRScs with a UK Biobank European ancestry linkage disequilibrium panel^36^. This method applies a continuous shrinkage prior to the SNP weights. PRScs was run in ‘auto’ mode on a per-chromosome basis. This mode places a standard half-Cauchy prior on the global shrinkage parameter and learns the global scaling parameter from the data; as a consequence, PRScs-auto does not require a validation data set for tuning. Based on the software default settings, only the 1.1 million SNPs found at HapMap3 sites that were also present in the UK Biobank were permitted to contribute to the score.

This score was applied to the entire UK Biobank. Participants related within 3 degrees of kinship to those who had undergone MRI, based on the precomputed relatedness matrix from the UK Biobank, were excluded from analysis^59^. We analyzed the relationship between this polygenic prediction of the LAmin and incident diagnoses of AF in the UK Biobank using a Cox proportional hazards model as implemented by the R *survival* package^65^. We excluded participants with disease that was diagnosed prior to enrollment in the UK Biobank. We counted survival as the number of years between enrollment and disease diagnosis (for those with disease) or until death, loss to follow-up, or end of follow-up time (for those without disease). We adjusted for covariates including sex, the cubic basis spline of age at enrollment, the interaction between the cubic basis spline of age at enrollment and sex, the genotyping array, the first five principal components of ancestry, and the cubic basis splines of height (cm), weight (kg), BMI (kg/m2), diastolic blood pressure (mmHg), and systolic blood pressure (mmHg).

### Definitions of diseases and medications

We defined AF or flutter, dilated cardiomyopathy, hypertrophic cardiomyopathy, heart failure, diabetes, and ischemic stroke based on self report, ICD codes, and procedural codes (**Supplementary Table 12**). The data were obtained from the UK Biobank in June 2020, at which time the recommended phenotype censoring date was March 31, 2020. The UK Biobank defines that date as the last day of the month for which the number of records is greater than 90% of the mean of the number of records for the previous three months (https://biobank.ndph.ox.ac.uk/ukb/exinfo.cgi?src=Data_providers_and_dates).

We identified participants taking antihypertensive medications based on the Anatomical Therapeutic Classification (ATC)^66^. Medications taken by UK Biobank participants were previously mapped to ATC codes^67^. We considered medications with ATC codes beginning with C02, C09, C08CA, C03AA, C08CA01, or C03BA04 to be antihypertensives (medication names enumerated in **Supplementary Table 13**).

## Data Availability

UK Biobank data are made available to researchers from research institutions with genuine research inquiries, following IRB and UK Biobank approval. GWAS summary statistics and polygenic score weights will be available upon publication at the Broad Institute Cardiovascular Disease Knowledge Portal ( http://www.broadcvdi.org ). LA measurements will be returned to the UK Biobank for use by any approved researcher. All other data are contained within the article and its supplementary information.

## Acknowledgments

We would like to thank Mary O’Reilly from the Broad Institute PATTERN Team for contributing to the graphical overview in Figure 1.

## Appendices

### Data availability

UK Biobank data are made available to researchers from research institutions with genuine research inquiries, following IRB and UK Biobank approval. GWAS summary statistics and polygenic score weights will be available upon publication at the Broad Institute Cardiovascular Disease Knowledge Portal (http://www.broadcvdi.org). LA measurements will be returned to the UK Biobank for use by any approved researcher. All other data are contained within the article and its supplementary information.

### Code availability

The code used to perform Poisson surface reconstruction from segmentation output is located at https://github.com/broadinstitute/ml4h and is available under an open-source BSD license.

### Author contributions

PTE and JPP conceived of the study. JPP annotated images. JPP trained the deep learning models. PD performed surface reconstruction. JPP, PD, SJ, and SHC conducted bioinformatic analyses. JPP, SHC, and PTE wrote the paper. All other authors contributed to the analysis plan or provided critical revisions.

### Sources of funding

This work was supported by the Fondation Leducq (14CVD01), and by grants from the National Institutes of Health to Dr. Ellinor (1RO1HL092577, K24HL105780) and Dr. Ho (R01HL134893, R01HL140224, K24HL153669). This work was supported by a John S LaDue Memorial Fellowship and the Sarnoff Cardiovascular Research Foundation Scholar Award to Dr. Pirruccello. Dr. Nauffal is supported by NIH grant 5T32HL007604-35. Dr. Khurshid is supported by NIH grant T32HL007208. Dr. Lubitz is supported by NIH grant 1R01HL139731 and American Heart Association 18SFRN34250007. This work was supported by a grant from the American Heart Association Strategically Focused Research Networks to Dr. Ellinor. Dr. Lindsay is supported by the Fredman Fellowship for Aortic Disease and the Toomey Fund for Aortic Dissection Research. This work was funded by a collaboration between the Broad Institute and IBM Research.

### Disclosures

Dr. Pirruccello has served as a consultant for Maze Therapeutics. Dr. Lubitz receives sponsored research support from Bristol Myers Squibb / Pfizer, Bayer AG, Boehringer Ingelheim, and Fitbit, and has consulted for Bristol Myers Squibb / Pfizer and Bayer AG, and participates in a research collaboration with IBM. Dr. Ng is employed by IBM Research. Dr. Ho is supported by a grant from Bayer AG focused on machine-learning and cardiovascular disease and a research grant from Gilead Sciences. Dr. Ho has received research supplies from EcoNugenics. Dr. Philippakis is employed as a Venture Partner at GV; he is also supported by a grant from Bayer AG to the Broad Institute focused on machine learning for clinical trial design. Dr. Ellinor is supported by a grant from Bayer AG to the Broad Institute focused on the genetics and therapeutics of cardiovascular diseases. Dr. Ellinor has also served on advisory boards or consulted for Bayer AG, Quest Diagnostics, MyoKardia and Novartis. Remaining authors report no disclosures.

