## Supplementary Note and Figures for "Deep Learning of Left Atrial Structure and Function Provides Link to Atrial Fibrillation Risk"

**Supplementary Note**	**1**

Semantic segmentation model training settings	2

Deep learning model quality assessment	3

Identification of abnormal atrial contraction	4

Atrial size is associated with AF, stroke, and heart failure	5

GWAS sensitivity analysis - LVEDV-indexing	6

Supplementary references	7

**Supplementary Figures**	**8**

Supplementary Figure 1 - Sample flow diagram	8

Supplementary Figure 2 - LV-adjusted left atrial phenotype Manhattan plots	9

Supplementary Figure 3 - Mendelian randomization method comparison plot for LAmin vs atrial fibrillation	10

Supplementary Figure 4 - Pleiotropic associations for variants used in Mendelian randomization	11

Supplementary Figure 5 - Mendelian randomization method comparison plot for LAmin vs atrial fibrillation after removing 3 pleiotropic variants	12

Supplementary Figure 6 - Mendelian randomization method comparison plot for atrial fibrillation vs LAmin	13

#### Semantic segmentation model training settings

The details of the hyperparameter settings of the semantic segmentation model for the short axis images is detailed in the **Online Methods**.

For the two-chamber long axis images, all images were resized initially to 104x92 pixels during the first half of training, and then to 208x186 pixels during the second half of training. The model was trained with a mini-batch size of 8 (with small images) or 4 (with large images). Maximum weight decay was 1E-03. Per-pixel cross entropy loss was minimized^1^. 30% of iterations were permitted to have an increasing learning rate during each epoch. When training with small images, the maximum learning rate was initially 1E-03, and training was performed over 30 epochs while keeping all weights frozen except for the final layer. When training with large images, the maximum learning rate was set to 1E-03, and the model was trained for 12 epochs while keeping all but the final layer frozen. Finally, all layers were unfrozen, the learning rate was decreased to 1E-06, and the model was retrained for an additional 8 epochs.

For the three-chamber long axis images, all images were resized initially to 128x128 pixels during the first half of training, and then to 256x256 pixels during the second half of training. The model was trained with a mini-batch size of 4 (with small images) or 2 (with large images). Maximum weight decay was 1E-02. Per-pixel cross entropy loss was minimized^1^. 30% of iterations were permitted to have an increasing learning rate during each epoch. When training with small images, the maximum learning rate was initially 1E-03, and training was performed over 20 epochs while keeping all weights frozen except for the final layer. Then, all layers were unfrozen, the learning rate was decreased to 3E-05, and the model was trained for an additional 20 epochs, with 80% of iterations permitted to have an increasing learning rate during each epoch. When training with large images, the maximum learning rate was set to 3E-04, and the model was trained for 15 epochs while keeping all but the final layer frozen; 20% of iterations were permitted to have an increasing learning rate during each epoch. Finally, all layers were unfrozen, the learning rate was decreased to 1E-07, and the model was retrained for an additional 7 epochs.

For the four-chamber long axis images, all images were resized initially to 76x104 pixels during the first half of training, and then to 150x208 pixels during the second half of training. The model was trained with a mini-batch size of 4 (with small images) or 2 (with large images). Maximum weight decay was 1E-02. Per-pixel cross entropy loss was minimized^1^. 30% of iterations were permitted to have an increasing learning rate during each epoch. When training with small images, the maximum learning rate was initially 1E-03, and training was performed over 50 epochs while keeping all weights frozen except for the final layer. Then, all layers were unfrozen, the learning rate was decreased to 3E-05, and the model was trained for an additional 15 epochs. When training with large images, the maximum learning rate was set to 3E-04, and the model was trained for 50 epochs while keeping all but the final layer frozen. Finally, all layers were unfrozen, the learning rate was decreased to 1E-07, and the model was retrained for an additional 15 epochs.

#### Deep learning model quality assessment

We used the Dice coefficient, which ranges from 0 for an image where no pixels overlap between human and machine labels, to 1 for an image with perfect overlap between human and machine labels, in order to assess the quality of automated semantic segmentation. In a held-out test set of 20 images from the two-chamber short axis view that were not used in training or validation, the average Dice coefficient was 0.89 (SD 0.06) for the left atrial blood pool. For 20 held-out images from the three-chamber view, the score was 0.88 (SD 0.07), and for 40 held-out images from the four-chamber view, the score was 0.94 (SD 0.03).

The short axis imaging sequence was not designed to capture the atria: the atrial short axis sequence was eliminated from the acquisition protocol to save time^2^. The left atrium was nevertheless recognizable in the basal-most segments of images obtained in the short axis view. In the short axis view, the average Dice score for the left atrium was 0.78 (SD 0.35) when weighted by the total number of pixels assigned to the left atrium by the cardiologist or the model, or 0.90 (SD 0.28) when considering images correctly identified by the model as having no left atrial pixels to have a Dice score of 1.

#### Identification of abnormal atrial contraction

In order to focus our analyses on participants unlikely to have LA measurements influenced by underlying pathology, we first sought to identify participants who appeared to have abnormal atrial contraction during the MRI. Although the MRI uses an electrocardiographic (ECG) signal for image acquisition, the underlying ECG signal is not available for analysis. Therefore, we trained a deep learning model to identify typical patterns of contraction based on volume throughout the cardiac cycle (**Online Methods**). Validation accuracy was 88.5%.

Among the 40,558 participants with LA volumes whose filling patterns could be analyzed, we identified 1,013 participants whose patterns either did not appear to be consistent with normal atrial contraction or could not be determined; of these, 376 (37%) had a pre-existing history of AF or atrial flutter. Of the 1,189 participants with a history of AF or atrial flutter, 376 (32%) had abnormal atrial contraction at the time of MRI. In contrast, of the 39,369 participants with no history of AF, only 637 (1.6%) had an abnormal atrial contraction pattern.

Among participants with no history of AF or atrial flutter, those with an abnormal atrial contraction had significantly elevated LA volumes (**Figure 2**; N = 637; LAmin: +1.3 standard deviations [SD], P = 3.1E-321; LAmax: +0.8 SD, P = 3.7E-103). The most extreme volumes were observed in participants with a history of AF or atrial flutter who had an abnormal atrial contraction pattern (N = 376; LAmin: +4.3 SD, P = 1.6E-1937; LAmax: +2.5 SD, P = 8.9E-623).

#### Atrial size is associated with AF, stroke, and heart failure

After excluding participants with abnormal atrial contraction patterns, we conducted analyses in the remaining 39,545 participants. First, we confirmed previous reports of the relationship between prevalent diseases and atrial size and function. Compared to the 38,732 UK Biobank participants without a diagnosis of AF or atrial flutter prior to MRI, the 813 with a pre-existing diagnosis had larger LA volumes (LAmin: +8.8mL, P = 9.2E-117; LAmax: +10.1mL, P = 1.5E-61) and a reduced LAEF (-4.6%, P = 9.7E-68). Participants with a history of heart failure or stroke also had elevated LA volumes (**Figure 3, left panel**; **Supplementary Table 1**).

We then examined the relationship between LA measurements and incident cardiovascular diseases. We excluded an additional 1,114 participants with prevalent AF, heart failure, or stroke diagnosed prior to MRI, and 1,525 with missing height, weight, or body mass index (BMI) measurements at the time of MRI. Only a brief period of follow-up time of 2.2 +/- 1.5 years after the MRI assessment center visit was available for most participants. Nevertheless, participants with a larger LA had a greater risk of subsequently being diagnosed with AF (293 incident AF diagnoses; hazard ratio [HR] 1.73 per standard deviation [SD] increase in LAmin; 95% CI 1.60-1.88; P = 4.0E-39; **Figure 3, right panel**). The LAmin was also associated with an increased risk of incident ischemic stroke (98 cases; HR 1.32 per SD; 95% CI 1.11-1.57; P = 2.0E-03) and heart failure (125 cases; HR 1.69; 95% CI 1.48-1.92; P = 1.3E-15). The associations between other LA measurements and these diseases are detailed in **Supplementary Table 2**.

We performed a sensitivity analysis that accounted for ECG features and left ventricular structure and function; this yielded a similar point estimate for LAmin as a marker of incident AF risk (HR 1.89 per SD; 95% CI 1.66-2.15; P = 4.5E-22). In this sensitivity analysis, LAmin remained a significant predictor of incident heart failure (HR 1.51 per SD; 95% CI 1.23-1.86; P = 8.1E-05) but not of incident ischemic stroke (HR 1.10 per SD; 95% CI 0.84-1.43; P = 0.48; **Supplementary Table 3**).

#### GWAS sensitivity analysis - LVEDV-indexing

Accounting for left ventricular volume can help to isolate genetic effects that are specific to the left atrium as well as to identify effects that are discordant between atrium and ventricle. However, adjusting for heritable covariates in GWAS can also induce associations via collider bias^3^. To attempt to identify LVEDV-indexed associations that were likely attributable to the adjustment for LVEDV, we tested each of the LVEDV-indexed lead SNPs for association with LVEDV.

We are not aware of a general solution to the interpretation of GWAS signals that incorporate adjustment for heritable covariates. However, we observed the LVEDV-indexed lead SNPs to fall into three patterns: first, some SNP associations appeared to be driven largely by the LVEDV adjustment rather than the LA volume. As an example of this pattern, the LVEDV-indexed LAmax association with *BAG3* (P=3.5E-10) was comparable to that for the LVEDV association with *BAG3* alone (P=2.1E-10), while the unadjusted LAmax measurement was not associated (P > 1E-3). At each of these loci, the effect direction in LVEDV was opposite to that in the respective LVEDV-indexed LA volume GWAS, which was expected. Practically, these signals appear to be driven by the LVEDV values, with the LA measurements acting as noise. Second, some SNP associations appeared to be driven by the LAmax association alone, with only minimal contribution from the LVEDV adjustment. For example, the LVEDV-indexed LAmax association with *IRAK1BP1* (P=2.0E-8) was similar to that for the LAmax association (P=2.7E-11), while the SNP was not associated with LVEDV (P > 1E-3). Third, some SNP associations appeared to be driven by the interplay between LA volumes and the LVEDV adjustment. For example, the *NEDD4L* locus was associated with LVEDV-indexed LAmax (P=4.7E-8) despite not being strongly associated with either LVEDV or LAmax alone (P > 1E-3 for both).

For the LVEDV-indexed LA volumes, 11 loci reached genome-wide significance for LAmax, 12 for LAmin, and four for LASV. Of these, six of the LVEDV-indexed LAmax loci had association P < 1E-3 with LVEDV, as did nine of the LAmin loci and two of the LASV loci. Novel loci that were not associated at genome-wide significance in the unadjusted GWAS, and which were not associated with LVEDV at 1E-3 or stronger, included *BLK*, *ANKRD1*, *MYH7*, and *NEDD4L* for LAmax; *CASQ2*, *DHX15*, *PROB1*, *UQCRB*, *ANKRD1*, and *MYH7* for LAmin, and *TNKS* and *HNRNPM* for LASV. Most of these loci were identified in the BSA-indexed GWAS.

### Supplementary Figures

#### Supplementary Figure 1 - Sample flow diagram

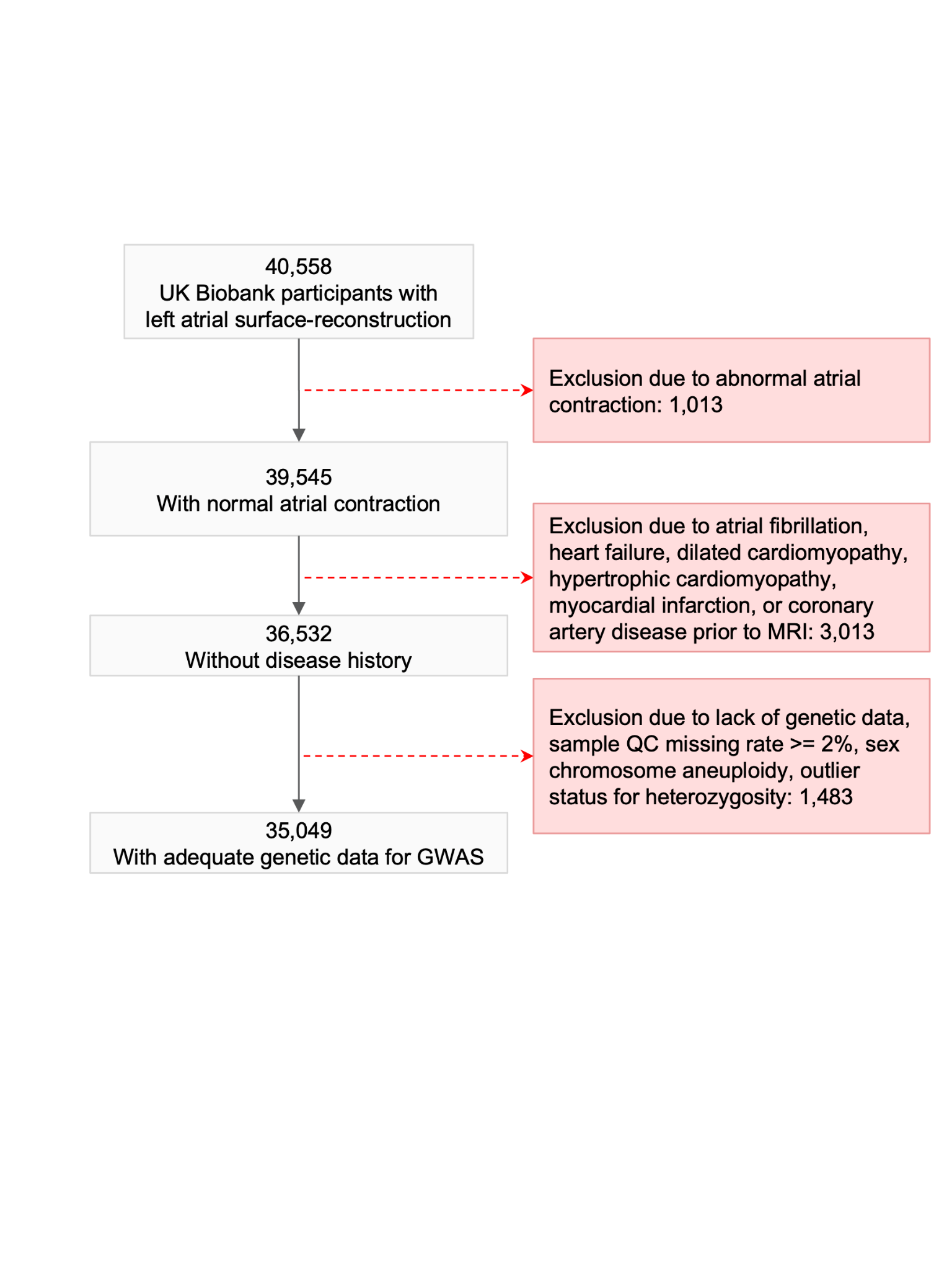

#### Supplementary Figure 2 - LV-adjusted left atrial phenotype Manhattan plots

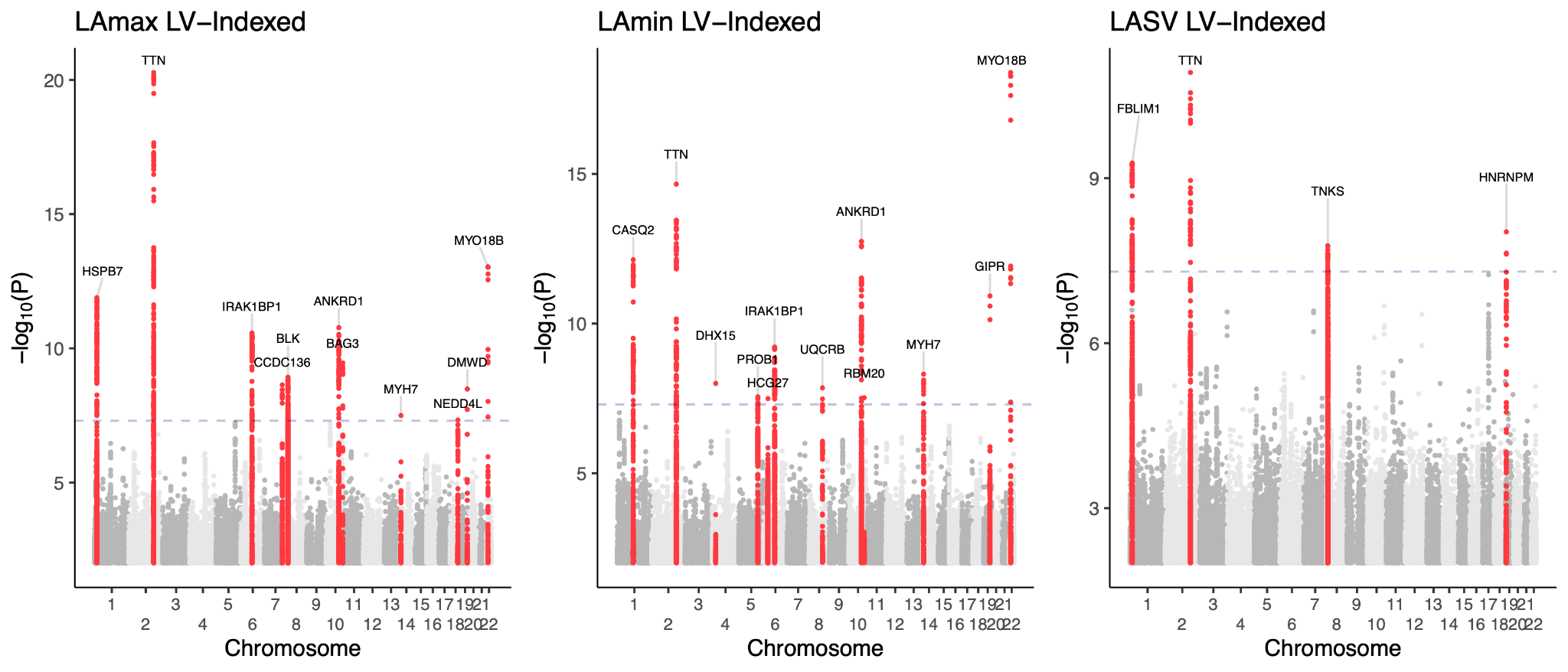

#### Supplementary Figure 3 - Mendelian randomization method comparison plot for LAmin vs atrial fibrillation

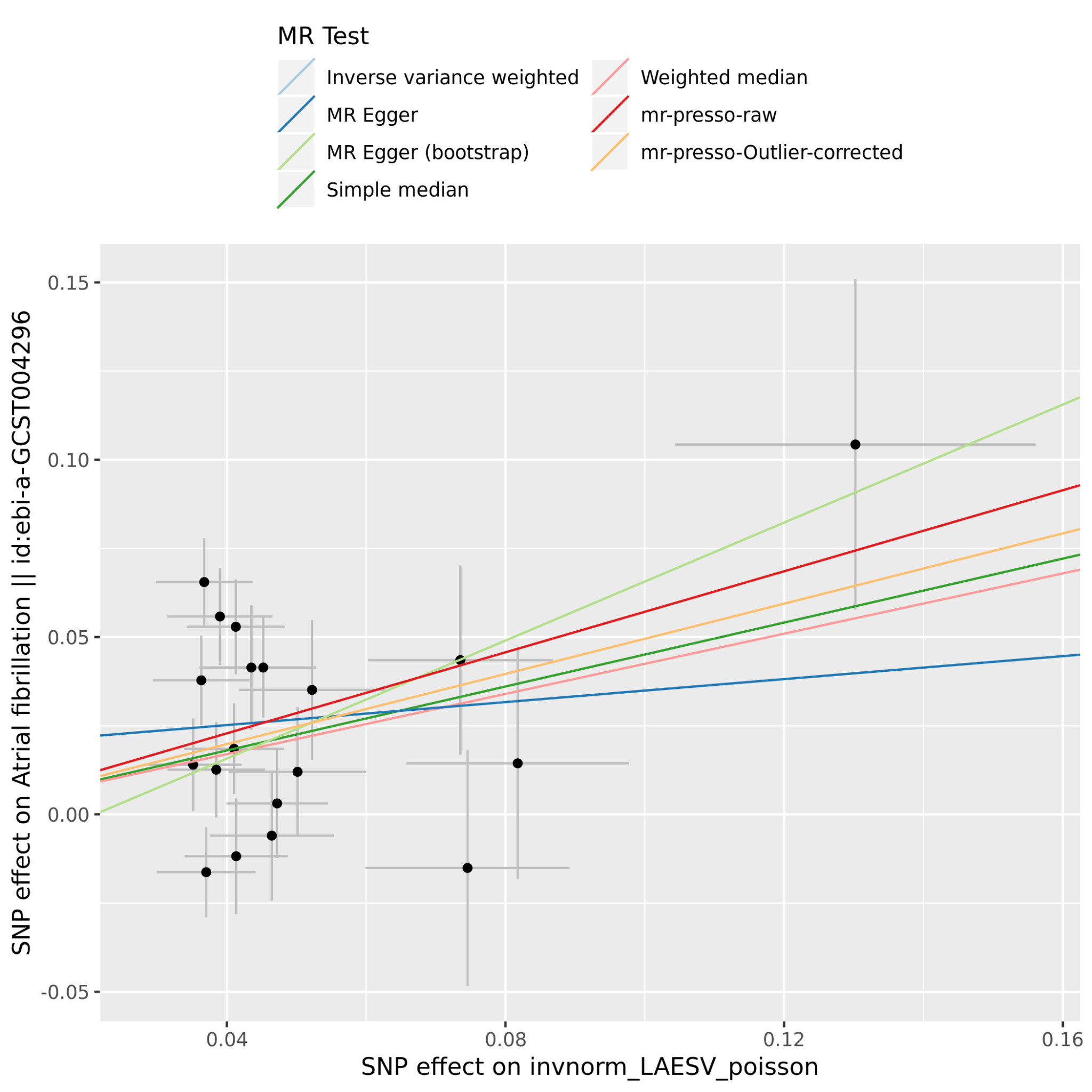

SNP effects on the exposure (X-axis) are plotted against SNP effects on the outcome (Y-axis). Here, the X-axis effect size comes from the LAmin volume GWAS in this manuscript (represented as “invnorm_LAESV_poisson”), while the Y-axis effect size comes from the Christophersen, et al, 2017 atrial fibrillation GWAS^4^.

#### Supplementary Figure 4 - Pleiotropic associations for variants used in Mendelian randomization

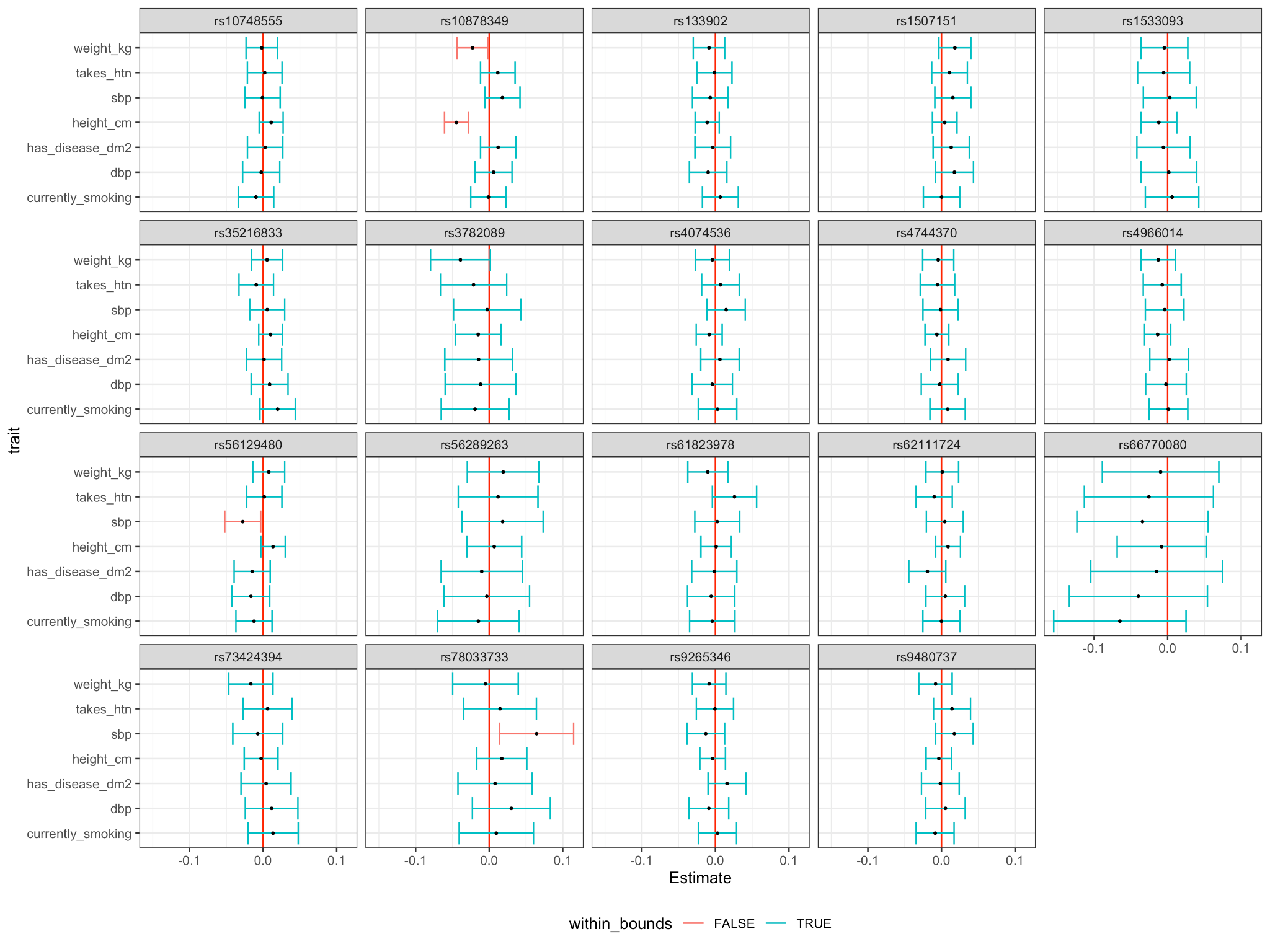

Each of the 19 SNPs from the LAmin Mendelian randomization analysis was tested for association with seven phenotypes previously identified as atrial fibrillation risk factors. For each SNP, this figure displays the point estimate of the effect of 1 unit change in the dosage of the non-reference allele on each trait. Traits where the association with the SNP achieves Bonferroni significance are shown in red. Three of the 19 SNPs were identified to have a significant association with at least one putative confounding factor (rs10878394 near *IRAK3*, rs56129480 near *SP3*, and rs78033733 near *MYL4*).

#### Supplementary Figure 5 - Mendelian randomization method comparison plot for LAmin vs atrial fibrillation after removing 3 pleiotropic variants

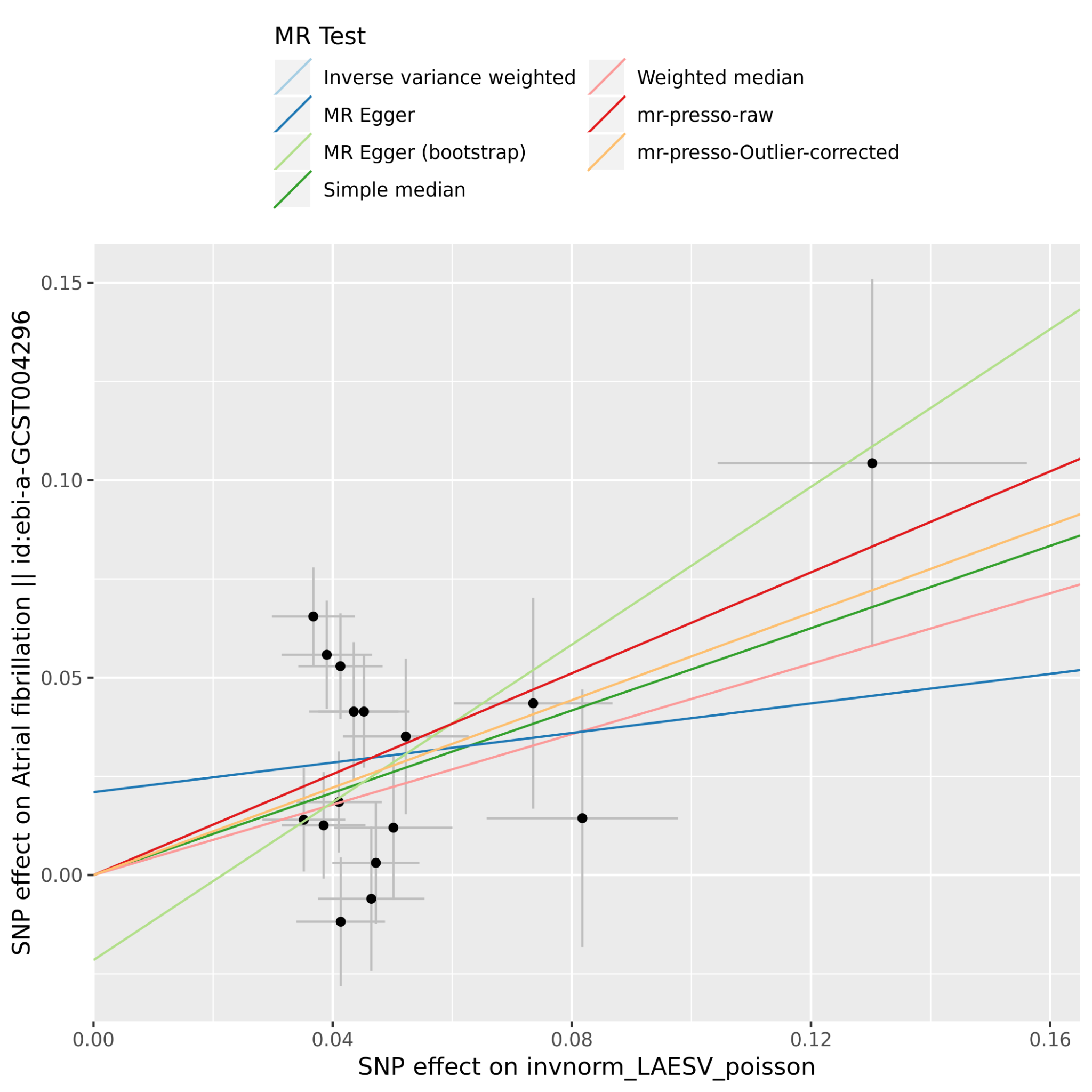

SNP effects on the exposure (X-axis) are plotted against SNP effects on the outcome (Y-axis). Here, the X-axis effect size comes from the LAmin volume GWAS in this manuscript (represented as “invnorm_LAESV_poisson”), while the Y-axis effect size comes from the Christophersen, et al, 2017 atrial fibrillation GWAS^4^.

#### Supplementary Figure 6 - Mendelian randomization method comparison plot for atrial fibrillation vs LAmin

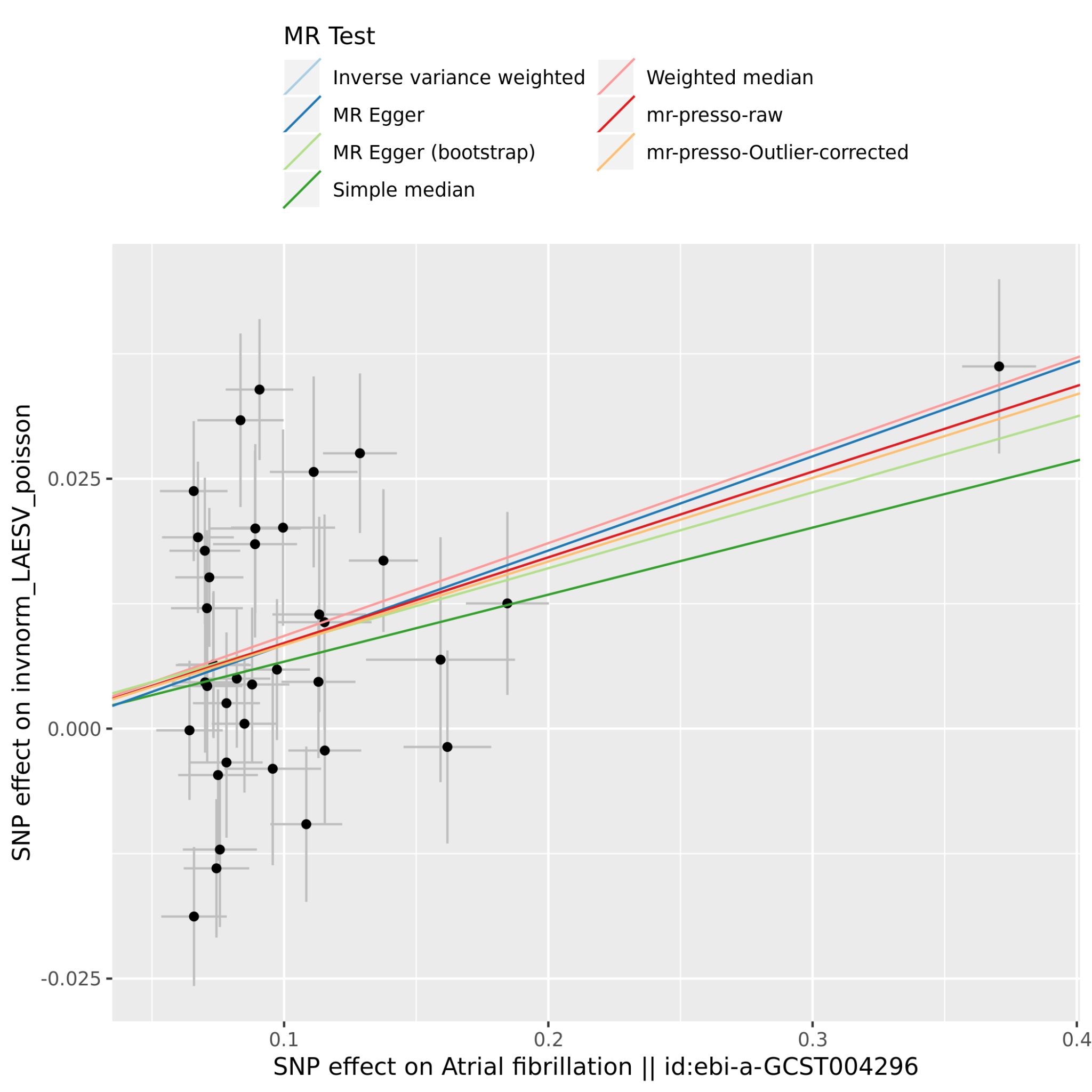

SNP effects on the exposure (X-axis) are plotted against SNP effects on the outcome (Y-axis). Here, the X-axis effect size comes from the Christophersen, et al, 2017 atrial fibrillation GWAS^4^, while the Y-axis effect size comes from the LAmin volume GWAS in this manuscript (represented as “invnorm_LAESV_poisson”).
